# Large Language Model Augmented Clinical Trial Screening

**DOI:** 10.1101/2024.08.27.24312646

**Authors:** Jacob Beattie, Dylan Owens, Ann Marie Navar, Luiza Giuliani Schmitt, Kimberly Taing, Sarah Neufeld, Daniel Yang, Christian Chukwuma, Ahmed Gul, Dong Soo Lee, Neil Desai, Dominic Moon, Jing Wang, Steve Jiang, Michael Dohopolski

## Abstract

**Purpose:** Identifying potential participants for clinical trials using traditional manual screening methods is time-consuming and expensive. Structured data in electronic health records (EHR) are often insufficient to capture trial inclusion and exclusion criteria adequately. Large language models (LLMs) offer the potential for improved participant screening by searching text notes in the EHR, but optimal deployment strategies remain unclear.

**Methods:** We evaluated the performance of GPT-3.5 and GPT-4 in screening a cohort of 74 patients (35 eligible, 39 ineligible) using EHR data, including progress notes, pathology reports, and imaging reports, for a phase 2 clinical trial in patients with head and neck cancer. Fourteen trial criteria were evaluated, including stage, histology, prior treatments, underlying conditions, functional status, etc. Manually annotated data served as the ground truth. We tested three prompting approaches (Structured Output (SO), Chain of Thought (CoT), and Self-Discover (SD)). SO and CoT were further tested using expert and LLM guidance (EG and LLM-G, respectively). Prompts were developed and refined using 10 patients from each cohort and then assessed on the remaining 54 patients. Each approach was assessed for accuracy, sensitivity, specificity, and micro F1 score. We explored two eligibility predictions: strict eligibility required meeting all criteria, while proportional eligibility used the proportion of criteria met. Screening time and cost were measured, and a failure analysis identified common misclassification issues.

**Results:** Fifty-four patients were evaluated (25 enrolled, 29 not enrolled). At the criterion level, GPT-3.5 showed a median accuracy of 0.761 (range: 0.554–0.910), with the Structured Output + EG approach performing best. GPT-4 demonstrated a median accuracy of 0.838 (range: 0.758–0.886), with the Self-Discover approach achieving the highest Youden Index of 0.729. For strict patient-level eligibility, GPT-3.5’s Structured Output + EG approach reached an accuracy of 0.611, while GPT-4’s CoT + EG achieved 0.65. Proportional eligibility performed better overall, with GPT-4’s CoT + LLM-G approach having the highest AUC (0.82) and Youden Index (0.60). Screening times ranged from 1.4 to 3 minutes per patient for GPT-3.5 and 7.9 to 12.4 minutes for GPT-4, with costs of $0.02–$0.03 for GPT-3.5 and $0.15–$0.27 for GPT-4.

**Conclusion:** LLMs can be used to identify specific clinical trial criteria but had difficulties identifying patients who met all criteria. Instead, using the proportion of criteria met to flag candidates for manual review maybe a more practical approach. LLM performance varies by prompt, with GPT-4 generally outperforming GPT-3.5, but at higher costs and longer processing times. LLMs should complement, not replace, manual chart reviews for matching patients to clinical trials.

## 1 Background

Clinical trials are essential for developing and validating new medical treatments. However, low patient accrual is a significant issue, with up to 20% of National Cancer Institute (NCI)-affiliated clinical trials failing due to insufficient enrollment [1]. This lack of participation reduces the predictive power of trials, leading to inconclusive results and significantly inflating research expenses[2, 3]. Factors impeding patient accrual include resource scarcity [4, 5], inefficient manual screening processes [6], and limited availability of research staff[7, 8]. Manual eligibility screening is particularly time-consuming, often requiring over 40 minutes per patient[6, 9]. Improving the efficiency of patient screening is, therefore, critical to ensure that clinical trials are sufficiently powered to produce meaningful results.

Leveraging electronic health records (EHR) offers a potential solution, but the necessary criteria for many trials are often buried in unstructured data, making it difficult to automate the screening process. Prior efforts to extract essential information from unstructured data have employed rule-based systems, traditional machine learning techniques, and natural language processing (NLP)[10–14]. These methods have significantly reduced screening times [12] and achieved high accuracy in specific tasks. However, implementing and customizing these techniques requires substantial clinical and technical expertise, as it demands a detailed knowledge of medical language and the ability to modify complex algorithms to accurately understand specific medical terms and conditions[11, 14, 15]. Large language models (LLMs) have recently emerged from advancements in NLP as powerful tools for analyzing unstructured text, demonstrating significant improvements in understanding complex language[16]. Recently, researchers have attempted to utilize LLMs to enhance various aspects of the clinical trial screening process. For example, LLMs have been used to expand training sets for other NLP models by generating reworded criteria descriptions [17] and to parse EHR to determine patient eligibility on a trial-level basis[18]. Previous work, including our own, demonstrated competitive performance in using LLMs to screen patient EHR on a criteria-level basis [19–21].

However, many questions remain about the optimal use of LLMs for clinical trial screening, including the best prompting approach, how it impacts model accuracy, and how different versions of LLMs compare. Additionally, it is important to determine how LLM-based screening can be integrated into clinical practice. To address these questions, we evaluated how basic and advanced prompting approaches influence the accuracy of screening patients both at the criteria level and overall.

## 2 Methods

### 2.1 Patients

We evaluated patient eligibility for a phase II trial investigating hypofractionated radiation therapy for head and neck cancer. A total of 14 criteria were identified based on trial inclusion/exclusion criteria for evaluation using the LLMs (Full clinical trial eligibility criteria are detailed in **Supplement A**). These criteria included: confirmed diagnosis of stage I-IVB squamous cell carcinoma of the oral cavity, oropharynx, hypopharynx, or larynx (”Carcinoma”); intermediate risk factors such as T3/4 disease, positive lymph nodes, close or positive margins, perineural invasion, or lymphovascular invasion (”Intermediate Risk Factors”); ECOG performance status of 0 to 2 (”ECOG *≥* 2”); no distant metastasis (”No Distant Metastasis”); no stage I or II glottic squamous cell carcinoma (”No Glottic Cancer”); no high-risk factors requiring concurrent chemotherapy (”No High-Risk Factors”); no additional non-skin primary cancers except for low/intermediate-risk prostate cancer and well-differentiated thyroid cancer (”No Synchronous Cancer”); no invasive malignancy with a disease-free interval of less than 3 years (”No Short Disease-Free Interval”); no prior radiotherapy overlapping with the current study’s radiation fields (”No Prior Radiotherapy”); no investigational agents for cancer treatment (”No Investigational Agents”); no uncontrolled intercurrent illnesses (”No Intercurrent Illness”); no pregnancy or nursing (”No Pregnancy”); no severe immunosuppression, including HIV or history of organ or stem cell transplants (”No Immunosuppression”); and no feeding tube dependence (”No Feeding Tube Dependence”). (**Table 1**. Two criteria were decided prior to any testing or fine-tuning to be excluded: age *≥*18 and use of contraception. The head and neck team does not commonly see patients *<* 18. Notes do not commonly report contraception use outside the setting of a clinical trial.

**Table 1.** Description of Criteria Used in the Study.

| **Criterion** | **Description** |
| --- | --- |
| **Carcinoma** | Patients must have a confirmed diagnosis of stage I-IVB squamous cell carcinoma of the oral cavity, oropharynx, hypopharynx, or larynx. |
| **Intermediate Risk Factors** | Includes T3/4 disease, positive lymph nodes, close or positive margins, perineural invasion, or lymphovascular invasion, indicating a moderate increase in the likelihood of cancer progression. |
| **ECOG $\leq 2$** | Patients must have an ECOG performance status of 0 to 2, indicating they are fully active or capable of self-care but unable to carry out any work activities. |
| **No Distant Metastasis** | The cancer must not have spread to distant organs or tissues beyond the original site. |
| **No Glottic Cancer** | Patients should not have stage I or II glottic squamous cell carcinoma, ensuring focus on other specified cancer locations. |
| **No High-Risk Factors** | Excludes patients with high-risk factors such as positive margins or extranodal extension requiring concurrent chemotherapy. |
| **No Synchronous Cancer** | Patients should not have additional non-skin primary cancers, except for low/intermediate-risk prostate cancer and well-differentiated thyroid cancer. |
| **No Short Disease-Free Interval** | Patients must not have had an invasive malignancy with a disease-free interval of less than 3 years. |
| **No Prior Radiotherapy** | Excludes patients with prior radiotherapy that overlaps with the current study's radiation fields, minimizing the risk of cumulative radiation effects. |
| **No Investigational Agents** | Patients should not be receiving any investigational agents for their cancer treatment during the study. |
| **No Intercurrent Illness** | Patients must not have uncontrolled intercurrent illnesses, such as infections or severe cardiac conditions, that could interfere with study participation. |
| **No Pregnancy** | Women must not be pregnant or nursing due to potential harm to the fetus or nursing infants from study interventions. |
| **No Immunosuppression** | Patients with severe immunosuppression, including those with HIV or a history of organ or stem cell transplants, are excluded to reduce infection risks. |
| **No Feeding Tube Dependence** | Patients must not be reliant on a feeding tube for nutrition, indicating an ability to maintain adequate nutrition orally. |

**Table 2.** Comparison of Per-Criterion Performance Metrics for GPT-3.5 and GPT-4 Using Different Prompting Approaches on the Testing Cohort (54 Patients: 25 Enrolled, 29 Not Enrolled). Abbreviations: Chain of Thought, CoT; Expert Guidance, EG; Large Language Model, LLM; LLM-Guidance, LLM-G; Custom Modules, CM; Accuracy, Acc; Sensitivity, Sens; Specificity, Spec; Youden Index, YI. The best approach measured using the YI: GPT-3.5, GPT-4.

| Model | Prompting Approach | Acc | Sens | Spec | Micro F1 | YI |
| --- | --- | --- | --- | --- | --- | --- |
| GPT-3.5 | Structured Output | 0.643 | 0.649 | 0.571 | 0.771 | 0.220 |
|  | Structured Output + LLM-G | 0.737 | 0.766 | 0.375 | 0.843 | 0.141 |
|  | Structured Output + EG | **0.910** | **0.953** | 0.375 | **0.951** | 0.328 |
|  | CoT | 0.644 | 0.636 | 0.750 | 0.768 | 0.386 |
|  | CoT + LLM-G | 0.554 | 0.537 | 0.768 | 0.691 | 0.305 |
|  | **CoT + EG** | 0.787 | 0.786 | **0.804** | 0.872 | **0.589** |
|  | Self-discover | 0.784 | 0.789 | 0.732 | 0.871 | 0.521 |
|  | Self-discover + CM | 0.820 | 0.827 | 0.732 | 0.895 | 0.559 |
| GPT-4 | Structured Output | 0.766 | 0.760 | **0.839** | 0.857 | 0.599 |
|  | Structured Output + LLM-G | 0.758 | 0.751 | **0.839** | 0.852 | 0.591 |
|  | Structured Output + EG | 0.872 | 0.876 | 0.821 | 0.927 | 0.697 |
|  | CoT | 0.775 | 0.773 | 0.804 | 0.864 | 0.576 |
|  | CoT + LLM-G | 0.804 | 0.801 | **0.839** | 0.883 | 0.641 |
|  | CoT + EG | 0.885 | **0.891** | 0.804 | **0.935** | 0.695 |
|  | **Self-discover** | **0.886** | 0.890 | **0.839** | **0.935** | **0.729** |
|  | Self-discover + CM | 0.884 | 0.889 | 0.821 | 0.934 | 0.710 |

Best statistic is listed in bolded for each evaluation category. The best-performing approach for the Youden index is listed in bold and highlighted in yellow.

Based on feedback from the study coordinators, three criteria were specifically identified as “high priority” due to their clinical complexity and importance: 1) “Carcinoma,” 2) “Intermediate Risk,” and 3) “No Short Disease-Free Interval.”

Our study sample included 35 patients who were enrolled in the trial and were considered “eligible”. We then randomly identified 40 patients. These patients were selected among new patients seen by the head and neck radiation oncology team. One patient was already enrolled in the trial and was excluded; therefore, 39 patients remained. Clinical research staff manually reviewed all patients’ EHRs to determine eligibility for each trial criterion.

The full clinical trial eligibility criteria are detailed in **Supplement A**.

### 2.2 Document Selection and Processing

We selected patient notes from surgical oncology, radiation oncology, and medical oncology, limited to documents from the last 6 months. Additionally, we included all pathology reports, imaging reports (limited to head and neck CT or MRI, and PET scans), and lab results pertinent to the study, allowing these reports to go back up to 1 year. We employed LlamaIndex as a data framework to organize and structure our patient data. LlamaIndex facilitated the ingestion and indexing of our medical records, creating an efficient searchable database. We used ChromaDB to create patient-specific vector database. This indexed structure enabled us to query the data effectively, allowing our retrieval system to select the 5 most relevant text passages. These passages then provided the necessary context for the LLM to determine if a patient met a particular criterion using these documents. These selected text passages were combined with the criteria-matching query and sent to an institutionally sanctioned, HIPAA-compliant Microsoft Azure Environment OpenAI API running GPT-3.5 (*API 0125*) or GPT-4 (*API 0125-Preview*) (**Figure 1**). The code was run on models available as of 12/2023.

**Fig. 1.**
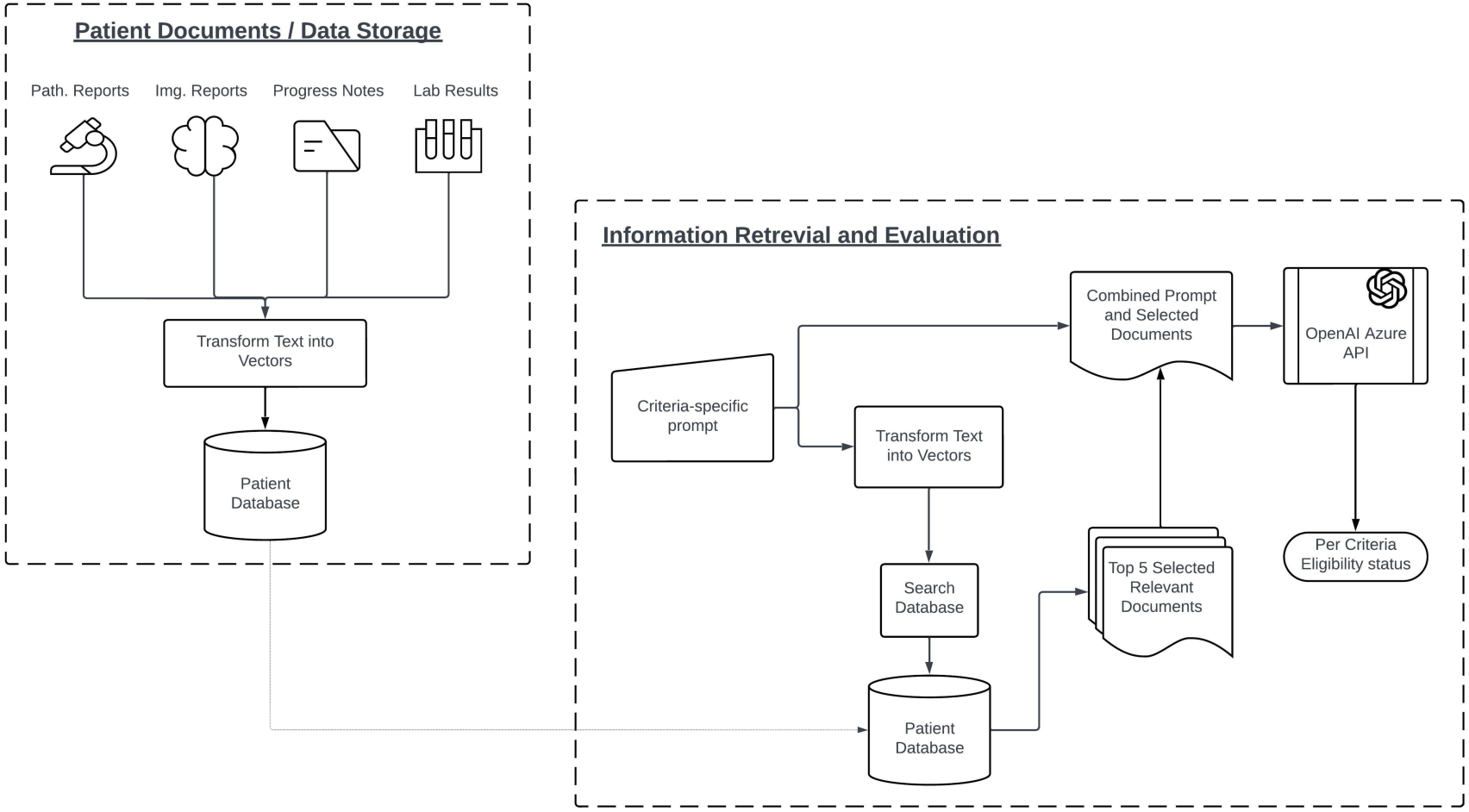
Overview of the document storage and retrieval process. Documents are used to create the vector database. Based on the criterion prompt, the 5 most relevant documents were compiled with the original prompt and sent to the LLM. The LLM provided an answer based on these selected documents. Abbreviation: Chain of Thought, CoT; Large Language Model, LLM.

### 2.3 Prompt and Guidance Generation

#### 2.3.1 Base Prompt

To develop the base prompt used in testing, we reserved 10 ineligible and 10 eligible patients (20 total) for error analysis-based prompt engineering. Initially, we created a foundational template using zero-shot prompting techniques[16]. We evaluated this initial prompt on the sample of 20 patients, identifying common reasoning failures.

After refining the base prompt, we tested three prompting approaches:

1. **Structured Output**: This prompting approach creates clear, organized responses that are easy to read and interpret, relying on the inherent reasoning of the LLM without any additional adjustments. See example in **Figure 2**.
2. **Chain-of-Thought (CoT)**[22]: Unlike Structured Output, CoT explicitly requests the model to think through problems step-by-step, explicitly articulating its reasoning process before reaching a conclusion. This approach breaks down complex tasks into intermediate steps, helping the model improve its problem-solving capabilities across various domains, including arithmetic, commonsense, and symbolic reasoning tasks. See example in **Figure 2**.
3. **Self-Discover**[23]: This prompting framework enhances the reasoning capabilities of LLMs for complex tasks, such as determining if a patient meets clinical trial criteria. It allows the LLM to autonomously construct and use logical reasoning structures. The model begins by selecting pre-made reasoning modules tailored to the task, such as analyzing patient data, comparing it to trial criteria, and drawing medical conclusions. The LLM then adapts these modules to create a customized reasoning process specific to the clinical trial evaluation. This structure guides the model through assessing patient eligibility by systematically considering factors like medical history, current health status, and trial requirements. Additionally, Custom Modules (CM) were designed by our group to further attempt to enhance the model’s performance in medical decision-making (Self-Discover + CM). See example in **Figure 2**.

**Fig. 2.**
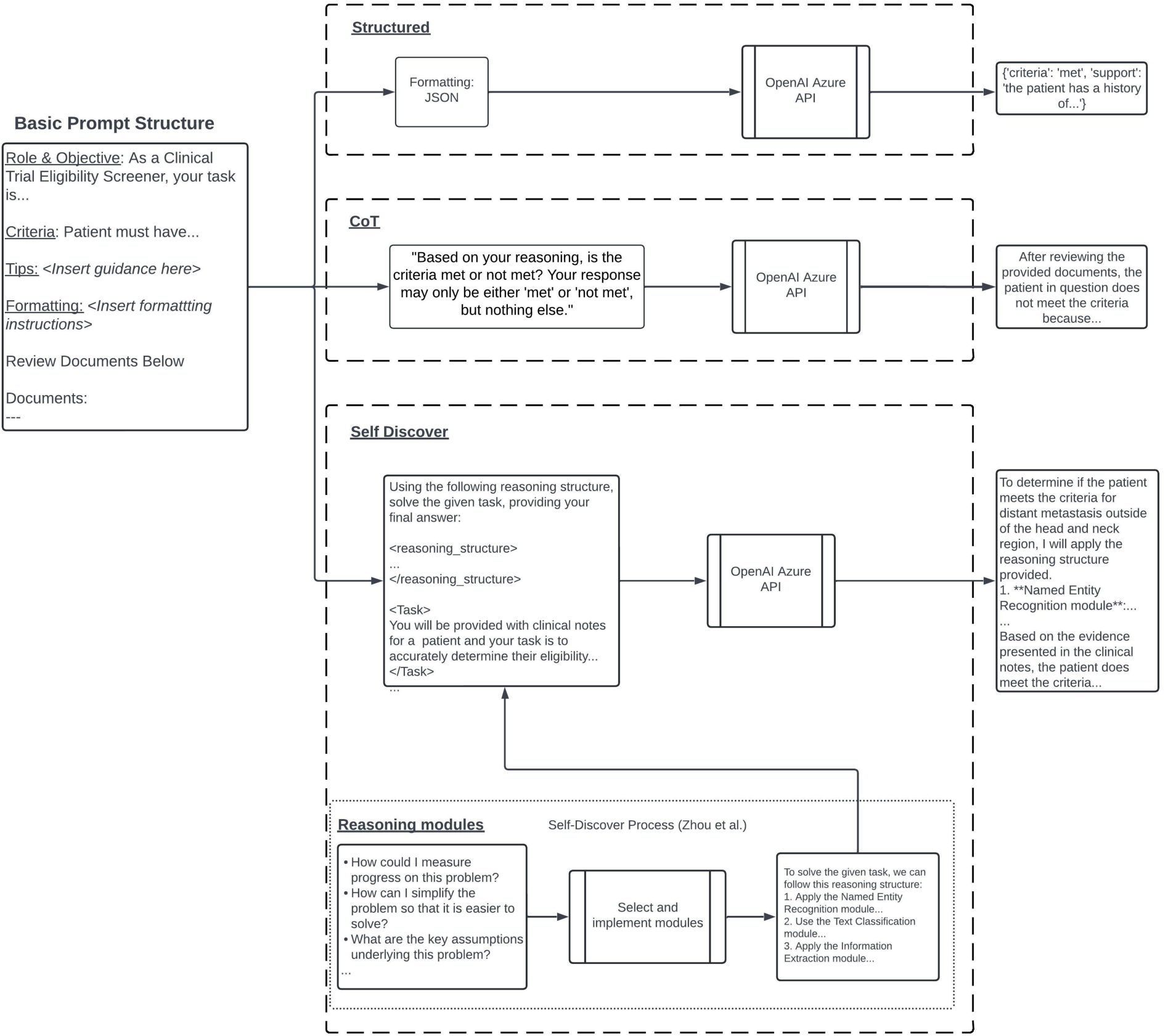
Depiction of prompting and response structure for structured output, CoT, Self-Discover. Self-Discover is designed to enhance the reasoning capabilities of LLMs. It allows LLMs to autonomously identify and construct logical reasoning structures to tackle complex tasks. The model first chooses from pre-made reasoning templates that best fit the task, such as determining if a patient meets a criterion. It then uses these templates to modify the original prompt, creating a cohesive and logical explanation. Abbreviation: Chain of Thought, CoT; Large Language Model, LLM.

**Fig. 3.**
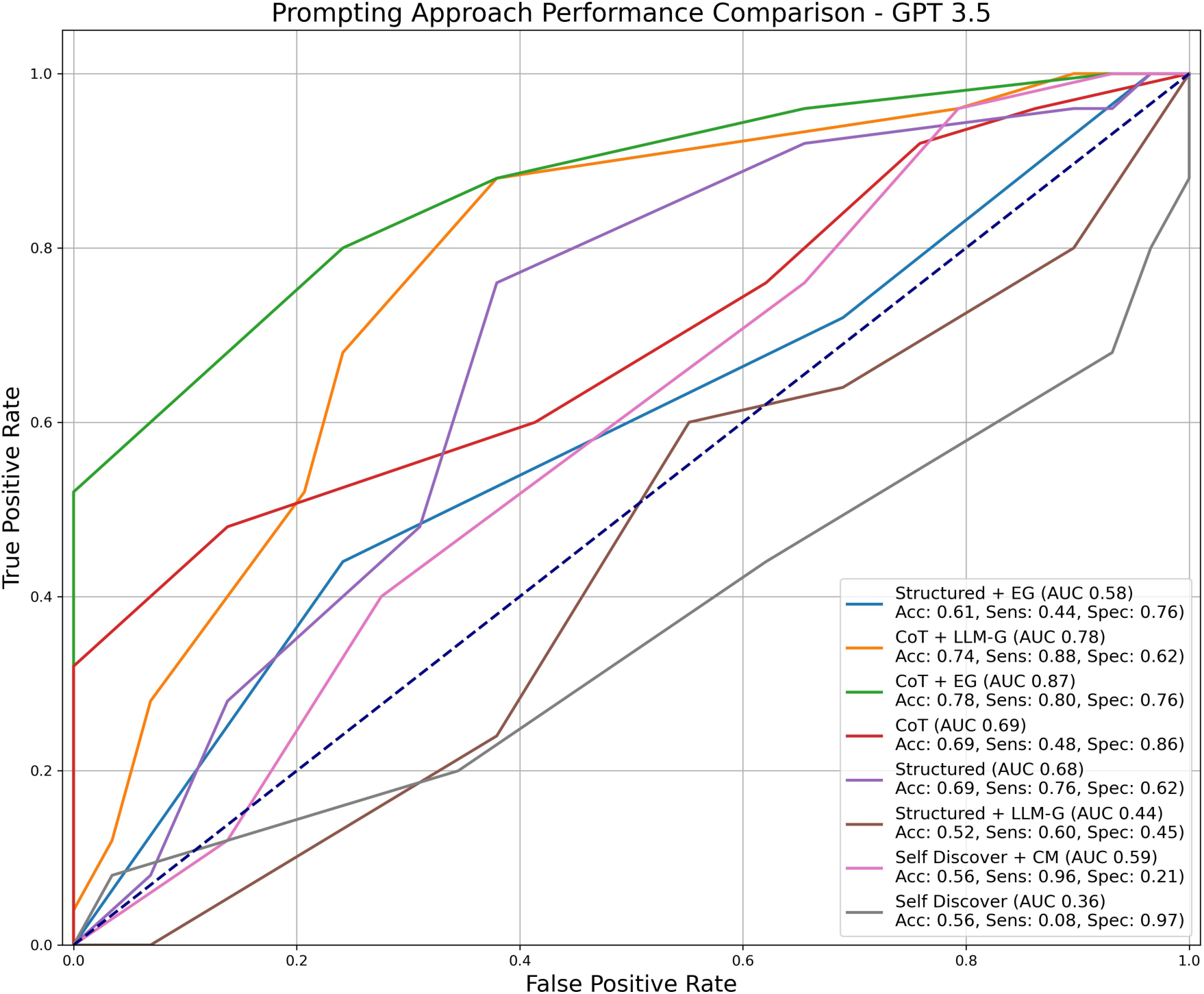
ROC-AUC Curves for prompting methods when using GPT-3.5. Proportional eligibility calculated the proportion of criteria met by each patient to derive a prediction score. For instance, if a patient met 8 out of 10 criteria, their prediction score would be 0.8. Since we had a proportion, we could use this as a predicted score. Using the predicted score generated by the proportional eligibility approach, we then created a Receiver Operating Characteristic (ROC) curve to assess prediction performance at different thresholds (i.e., % criteria needed to be met to be classified as eligible). Acc, Sens, and Spec were calculated using the threshold that maximized the Youden Index. Abbreviations: Chain of Thought, CoT; Expert Guidance, EG; Custom Modules, CM; Accuracy, Acc; Sensitivity, Sens; Specificity, Spec.

**Fig. 4.**
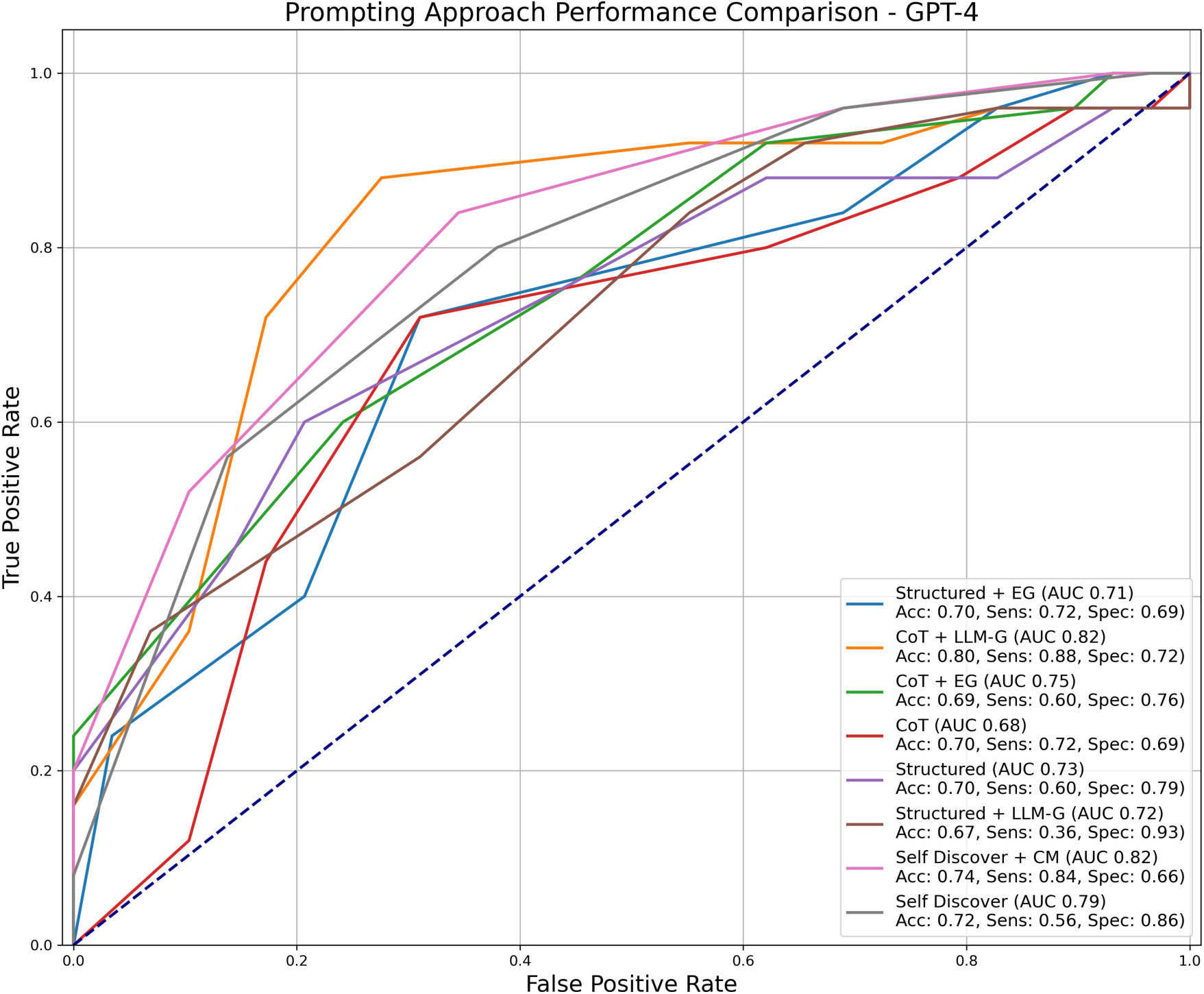
ROC-AUC Curves for prompting methods when using GPT-4. Proportional eligibility calculated the proportion of criteria met by each patient to derive a prediction score. For instance, if a patient met 8 out of 10 criteria, their prediction score would be 0.8. Since we had a proportion, we could use this as a predicted score. Using the predicted score generated by the proportional eligibility approach, we then created a Receiver Operating Characteristic (ROC) curve to assess prediction performance at different thresholds (i.e., % criteria needed to be met to be classified as eligible). Acc, Sens, and Spec were calculated using the threshold that maximized the Youden Index. Abbreviations: Chain of Thought, CoT; Expert Guidance, EG; Custom Modules, CM; Accuracy, Acc; Sensitivity, Sens; Specificity, Spec.

#### 2.3.2 Guidance

We applied different types of guidance to enhance the performance of the Structured Output and CoT prompting approaches. Guidance was not applied to the Self-Discover approach, as we wanted to stay true to its autonomous nature and explore its effectiveness without external inputs.

- **No Guidance**: No additional guidance was provided.
- **Expert Guidance (EG)**: A physician provided a reasoning framework and relevant keywords and phrases to enhance the document retrieval and reasoning processes. These tips were iteratively tested and refined on the same cohort used to create the base prompt (10 eligible and 10 ineligible).
- **LLM-Guidance (LLM-G)**: The LLM provided its own guidance, which included relevant keywords and phrases.

Detailed prompts for each method are provided in **Supplement A**.

### 2.4 Analyses

We conducted several analyses to evaluate the performance of different prompting approaches using both GPT-3.5 and GPT-4. Our analyses included accuracy, sensitivity, specificity, Youden Index (sensitivity + specificity - 1), micro F1 score, and ROC AUC. These metrics were calculated on a per-criterion and per-patient basis where applicable. The testing cohort consisted of 54 patients, separate from the patients used for base prompt engineering and EG generation.

#### 2.4.1 Identifying Best-Performing Methods for GPT-3.5 and GPT-4 at the Criterion Level

We compared all three prompting approaches—Structured Output, CoT, and Self-Discover. Guidance (EG or LLM-G) was applied only to the Structured Output and CoT approaches to enhance their performance. Guidance was not applied to the Self-Discover approach, as we aimed to maintain its fully autonomous nature and evaluate its effectiveness without external inputs.

We conducted sub-analyses on three selected criteria: Carcinoma, Intermediate-Risk, and No Short Disease-Free Interval. These criteria were chosen due to their clinical complexity and balanced distribution of criteria met versus unmet, providing a robust test of the models’ capabilities.

#### 2.4.2 Assessing Patient-Level Eligibility for Trial Enrollment

To evaluate the models’ performance in making patient-level predictions for trial enrollment, we used two approaches: strict eligibility and proportional eligibility.

Strict eligibility classified patients as “eligible” only if they met all criteria. We calculated accuracy, sensitivity, and specificity for this approach. Proportional eligibility, on the other hand, calculated the proportion of criteria met by each patient to derive a prediction score. For instance, if a patient met 8 out of 10 criteria, their prediction score would be 0.8. Since we had a proportion, we could use this as a predicted score. Using the predicted score generated by the proportional eligibility approach, we then created a Receiver Operating Characteristic (ROC) curve to assess prediction performance at different thresholds (i.e., % criteria needed to be met to be classified as eligible). Accuracy, sensitivity, and specificity were calculated using the threshold that maximized the Youden Index.

#### 2.4.3 Screening Time and Cost

For each approach, We measured the average screening time and cost using a sample of 5 randomly selected patients. Microsoft Azure’s OpenAI model pricing is based on the amount of text processed (EHR documents) and generated (LLM response). In this context, text is measured in units called tokens, where a token can be as short as one character or as long as one word. We counted the average number of tokens per prompt (input text) and response (output text) for the selected patients and applied the pricing models for GPT-3.5 and GPT-4 to calculate the average price.

#### 2.4.4 Failure Analysis

After final prompt refinement, we analyzed a random subset of 42 misclassifications (21 errors each for GPT-3.5 and GPT-4) in the testing dataset (54 patients: 25 eligible and 29 ineligible) across prompting approaches to identify common failure types. We categorized the errors into two main types:

- **Incorrect Understanding**: This occurred when the pipeline selected the appropriate text but came to the wrong conclusion. For example, the model might correctly identify the location of a tumor but incorrectly determine its stage.
- **Missing Information**: This happened when the model was unable to identify the correct text in the patient chart, even though it was present. For instance, the model might fail to locate a specific date or clinical note that was crucial for the criteria assessment.

## 3 Results

### 3.1 Best-Performing Approaches at the Criterion Level for GPT-3.5 and GPT-4

#### 3.1.1 Summary of Overall Performance

For GPT-3.5, accuracy was 0.761 (range: 0.554–0.910), sensitivity was 0.776 (range: 0.537–0.953), specificity was 0.732 (range: 0.375–0.804), and the Youden Index was 0.357 (range: 0.141–0.589). The Structured Output + EG approach achieved the highest accuracy at 0.910 and the highest sensitivity at 0.953. However, the best-performing approach in terms of the Youden Index was CoT + EG, with a YI of 0.589, balancing a sensitivity of 0.786 and a specificity of 0.804.

For GPT-4, accuracy was 0.838 (range: 0.758–0.886), sensitivity was 0.839 (range: 0.751–0.891), specificity was 0.830 (range: 0.804–0.839), and the Youden Index was 0.668 (range: 0.576–0.729). The Self-Discover approach performed the best with a Youden Index of 0.729, showing a strong balance between sensitivity (0.890) and specificity (0.839). ACoT + EG achieved similar performance, accuracy of 0.885 and a YI of 0.695. The Self-Discover + CM approach also performed well, with a YI of 0.710.

#### 3.1.2 Detailed Youden Index Comparisons

In the Structured Output approach for GPT-3.5, the Youden Index increased from 0.220 to 0.328 with EG but decreased to 0.141 with LLM-G. For GPT-4, the Youden Index improved from 0.599 to 0.697 with EG and was slightly lower at 0.591 with LLM-G.

In the CoT approach for GPT-3.5, the Youden Index increased from 0.386 to 0.589 with EG but decreased to 0.305 with LLM-G. For GPT-4, the Youden Index increased from 0.576 to 0.695 with EG and to 0.641 with LLM-G.

In the Self-Discover approach, GPT-4 outperformed GPT-3.5. The Youden Index for GPT-4 was 0.729 with the standard Self-Discover method, compared to 0.521 for GPT-3.5. Even with the addition of Custom Modules (CM), GPT-4 continued to show superior performance, achieving a Youden Index of 0.710 versus 0.559 for GPT-3.5.

### 3.2 Clinically Complex Clinical Criteria Evaluation

#### 3.2.1 Carcinoma Criterion

For GPT-3.5, median accuracy was 0.870 (range: 0.741–0.889), sensitivity was 0.886 (range: 0.629–0.971), specificity was 0.842 (range: 0.316–1.000), and the Youden Index was 0.629 (range: 0.287–0.800). The highest accuracy (0.889) was achieved with the Self-Discover +/- CM approach, while the highest Youden Index (0.800) was observed with the CoT approach.

For GPT-4, median accuracy was 0.944 (range: 0.944–0.963), sensitivity was 0.971 (consistent), specificity was 0.895 (range: 0.895–0.947), and the Youden Index was 0.866 (range: 0.866–0.919). The highest accuracy (0.963) and Youden Index (0.919) were achieved with the Structured Output and CoT + LLM-G approaches.

#### 3.2.2 Intermediate-Risk Criterion

For GPT-3.5, median accuracy was 0.722 (range: 0.537–0.815), sensitivity was 0.684 (range: 0.342–0.921), specificity was 0.750 (range: 0.375–1.000), and the Youden Index was 0.438 (range: 0.263–0.684). The highest accuracy (0.815) was achieved with the Structured Output + EG approach, and the highest Youden Index (0.684) was observed with the CoT + EG approach.

For GPT-4, median accuracy was 0.852 (range: 0.833–0.870), sensitivity was 0.868 (range: 0.842–0.895), specificity was 0.812 (range: 0.750–0.875), and the Youden Index was 0.645 (range: 0.618–0.743). The highest accuracy (0.870) was achieved with the Structured Output + EG, Self-Discover, and Self-Discover + CM approaches, while the highest Youden Index (0.743) was observed with the two Self-Discover approaches.

#### 3.2.3 No Short Disease-Free Interval Criterion

For GPT-3.5, median accuracy was 0.741 (range: 0.556–0.870), sensitivity was 0.816 (range: 0.571–0.918), specificity was 0.400 (range: 0.000–0.800), and the Youden Index was 0.118 (range: - 0.286–0.657). The highest accuracy (0.870) was achieved with the CoT + EG approach, while the highest Youden Index (0.657) was observed with the CoT + LLM-G approach.

For GPT-4, median accuracy was 0.574 (range: 0.167–0.704), sensitivity was 0.551 (range: 0.082–0.694), specificity was 0.800 (range: 0.600–1.000), and the Youden Index was 0.351 (range: 0.082–0.571). The highest sensitivity (0.571) and Youden Index (0.571) were achieved with the Structured Output + EG approach, while the Self-Discover approach provided balanced performance with an accuracy of 0.630 and a Youden Index of 0.412.

Performance on all criteria for all methods can be found in **Table 3**.

**Table 3:** Comparison of Criterion-Level Performance Metrics for GPT-3.5 and GPT-4 on Selected Criteria (Carcinoma, Intermediate-Risk, No Short Disease-Free Interval) Using Different Prompting Approaches. Abbreviations: Chain of Thought, CoT; Expert Guidance, EG; Custom Modules, CM; Accuracy, Acc; Sensitivity, Sens; Specificity, Spec; Youden Index, YI. ^*^Guidance (EG and LLM-G) was not provided in the Self-Discover approach due to its inherent design, which relies on the model’s ability to autonomously identify appropriate reasoning modules and adjust the prompt accordingly. The best approach measured using YI: GPT-3.5, GPT-4, Both.

| Criterion | Approach | Acc (GPT-3.5/4) | Sens (GPT-3.5/4) | Spec (GPT-3.5/4) | YI (GPT-3.5/4) |
| --- | --- | --- | --- | --- | --- |
| Carcinoma | **Structured Output** | 0.778 / 0.963 | 0.971 / 0.971 | 0.421 / 0.947 | 0.392 / 0.919 |
|  | Structured Output + LLM-G | 0.741 / 0.944 | 0.971 / 0.971 | 0.316 / 0.895 | 0.287 / 0.866 |
|  | Structured Output + EG | 0.759 / 0.944 | 0.971 / 0.971 | 0.368 / 0.895 | 0.340 / 0.866 |
|  | **CoT** | 0.870 / 0.944 | 0.800 / 0.971 | 1.000 / 0.895 | 0.800 / 0.866 |
|  | **CoT + LLM-G** | 0.870 / 0.963 | 0.886 / 0.971 | 0.842 / 0.947 | 0.728 / 0.919 |
|  | CoT + EG | 0.759 / 0.944 | 0.629 / 0.971 | 1.000 / 0.895 | 0.629 / 0.866 |
|  | **Self-Discover** | 0.889 / 0.944 | 0.886 / 0.971 | 0.895 / 0.895 | 0.780 / 0.866 |
|  | Self-Discover + CM | 0.889 / 0.944 | 0.914 / 0.971 | 0.842 / 0.895 | 0.756 / 0.866 |
| Intermediate-Risk | Structured Output | 0.815 / 0.833 | 0.868 / 0.842 | 0.688 / 0.812 | 0.556 / 0.655 |
|  | Structured Output + LLM-G | 0.759 / 0.852 | 0.921 / 0.895 | 0.375 / 0.750 | 0.296 / 0.645 |
|  | Structured Output + EG | 0.685 / 0.870 | 0.763 / 0.895 | 0.500 / 0.812 | 0.263 / 0.707 |
|  | CoT | 0.722 / 0.833 | 0.658 / 0.868 | 0.875 / 0.750 | 0.533 / 0.618 |
|  | CoT + LLM-G | 0.667 / 0.833 | 0.632 / 0.868 | 0.750 / 0.750 | 0.382 / 0.618 |
|  | **CoT + EG** | 0.778 / 0.852 | 0.684 / 0.895 | 1.000 / 0.750 | 0.684 / 0.645 |
|  | **Self-Discover** | 0.537 / 0.870 | 0.342 / 0.868 | 1.000 / 0.875 | 0.342 / 0.743 |
|  | **Self-Discover + CM** | 0.630 / 0.870 | 0.500 / 0.868 | 0.938 / 0.875 | 0.438 / 0.743 |
| No Short<br>Disease-Free Interval | Structured Output | 0.556 / 0.389 | 0.571 / 0.347 | 0.400 / 0.800 | -0.029 / 0.147 |

Table 3 – continued from previous page
| Criterion | Approach | Acc (GPT-3.5/4) | Sens (GPT-3.5/4) | Spec (GPT-3.5/4) | YI (GPT-3.5/4) |
| --- | --- | --- | --- | --- | --- |
|  | Structured Output + LLM-G | 0.667 / 0.167 | 0.694 / 0.082 | 0.400 / 1.000 | 0.094 / 0.082 |
|  | **Structured Output + EG** | 0.815 / 0.611 | 0.857 / 0.571 | 0.400 / 1.000 | 0.257 / 0.571 |
|  | CoT | 0.741 / 0.574 | 0.816 / 0.551 | 0.000 / 0.800 | -0.184 / 0.351 |
|  | **CoT + LLM-G** | 0.852 / 0.389 | 0.857 / 0.347 | 0.800 / 0.800 | 0.657 / 0.147 |
|  | CoT + EG | 0.870 / 0.704 | 0.918 / 0.694 | 0.400 / 0.800 | 0.318 / 0.494 |
|  | Self-Discover | 0.648 / 0.630 | 0.714 / 0.612 | 0.000 / 0.800 | -0.286 / 0.412 |
|  | Self-Discover + CM | 0.852 / 0.556 | 0.918 / 0.551 | 0.200 / 0.600 | 0.118 / 0.151 |

### 3.3 Assessing Patient-Level Eligibility for Trial Enrollment

#### 3.3.1 Strict Eligibility

For GPT-3.5, the median accuracy was 0.54 (range: 0.50–0.61), with the highest accuracy in the Structured + EG approach at 0.611. The median sensitivity was 0.00 (range: 0.00–0.44), with the Structured + EG approach reaching the highest sensitivity at 0.44. The median specificity was 1.00 (range: 0.76–1.00), with multiple approaches, including CoT + LLM-G, CoT + EG, CoT, Structured, Self Discover + CM, and Self Discover, achieving the highest specificity at 1.00. The median Youden Index was 0.00 (range: -0.07–0.20), with the Structured + EG approach achieving the highest Youden Index at 0.20.

For GPT-4, the median accuracy was 0.61 (range: 0.54–0.65), with the highest accuracy in the CoT + EG approach at 0.65. The median sensitivity was 0.16 (range: 0.00–0.24), with the CoT + EG and Structured + EG approaches both reaching the highest sensitivity at 0.24. The median specificity was 1.00 (range: 0.97–1.00), with multiple approaches, including CoT + LLM-G, CoT + EG, CoT, Structured, Structured + LLM-G, Self Discover + CM, and Self Discover, achieving the highest specificity at 1.00. The median Youden Index was 0.20 (range: 0.00–0.24), with the CoT + EG approach achieving the highest Youden Index at 0.24 (**Table C3**).

#### 3.3.2 Proportional Eligibility

For GPT-3.5, the median AUC was 0.64 (range: 0.36–0.87), with the CoT + EG approach having the highest AUC at 0.87. The median accuracy was 0.65 (range: 0.52–0.78), with the highest accuracy observed in the CoT + EG approach at 0.78. The median sensitivity was 0.68 (range: 0.08–0.96), where the Self Discover + CM approach had the highest sensitivity at 0.96. The median specificity was 0.69 (range: 0.21–0.97), with the Self Discover approach showing the highest specificity at 0.97. The median Youden Index was 0.27 (range: 0.05–0.56), with the CoT + EG approach achieving the highest Youden Index at 0.56.

For GPT-4, the median AUC was 0.74 (range: 0.68–0.82), with the CoT + LLM-G approach having the highest AUC at 0.82. The median accuracy was 0.70 (range: 0.67–0.80), with the highest accuracy observed in the CoT + LLM-G approach at 0.80. The median sensitivity was 0.66 (range: 0.36–0.88), where the CoT + LLM-G approach also had the highest sensitivity at 0.88. The median specificity was 0.74 (range: 0.66–0.93), with the Structured + LLM-G approach reaching the highest specificity at 0.93. The median Youden Index was 0.41 (range: 0.29–0.60), with the CoT + LLM-G approach achieving the highest Youden Index at 0.60.

### 3.4 Screening Time and Cost

Screening a single patient using GPT-3.5 took between 1.4 and 3 minutes, while using GPT-4 took between 7.9 and 12.4 minutes. The longer screening times with GPT-4 are likely due to the additional computing power required for this more advanced model within Microsoft Azure’s cloud service. The cost of screening a single patient ranged from $0.02 to $0.03 with GPT-3.5 and from $0.15 to $0.27 with GPT-4.

**Table 4** details the full results for screening time and cost across different prompting approaches.

**Table 4.** Screening Time and Cost for Each Approach. Microsoft Azure’s OpenAI model pricing is based on the amount of text processed (EHR documents) and generated (LLM response). Text is measured in units called tokens, where a token can be as short as one character or as long as one word. We counted the average number of tokens per prompt (input text) and response (output text) for the selected patients and applied the pricing models for GPT-3.5 and GPT-4 to calculate the average price. Guidance was added to calculate maximum cost (increased input tokens). Abbreviations: Abbreviations: Chain of Thought, CoT; Expert Guidance, EG; Large Language Model, LLM; LLM-Guidance, LLM-G.

| Model | Prompting Approach | Avg. Screening Time (min.) | Single Patient Cost (USD) |
| --- | --- | --- | --- |
| GPT-3.5 | Structured Output + EG | 1.4 | \$0.02 |
| | CoT + EG | 1.8 | \$0.02 |
| | Self-discover | 3.0 | \$0.03 |
| GPT-4 | Structured Output + EG | 7.9 | \$0.15 |
| | CoT + EG | 9.5 | \$0.22 |
| | Self-discover | 12.4 | \$0.27 |

### 3.5 Failure Analysis

Upon analyzing 42 misclassifications—21 errors made by GPT-4 and 21 by GPT-3.5—across various prompt structures, we observed two prominent failure types:

1. **Incorrect Understanding**: For GPT-4, 20 out of 21 errors (95%) were due to incorrect understanding. For GPT-3.5, 15 out of 21 errors (71%) were attributable to incorrect understanding. Overall, 35 of the 42 misclassifications (83%) involved situations where the model had the correct text but made the wrong conclusion, often failing to correctly interpret record dates, tumor locations, or criteria requirements.
2. **Missing Information**: Missing information was a more significant issue for GPT-3.5, with 6 out of 21 errors (29%) attributed to this problem. For GPT-4, only 1 out of 21 errors (5%) was due to missing information. Overall, 7 of the 42 misclassifications (17%) occurred because the model did not select the appropriate text needed to answer the question correctly, leading to a lack of relevant evidence.

## 4 Discussion

This study evaluated the use of LLMs for patient screening in clinical trials. While GPT-3.5 and GPT-4 effectively identified likely eligible patients, they struggled to correctly identify all criteria for each patient. Using the proportion of criteria met was a more effective method for identifying potential candidates. Performance varied across prompting methods: GPT-3.5 performed better with clear instructions but was less effective with advanced, fully automated approaches like Self-Discover. In contrast, GPT-4 performed well across all approaches, including Self-Discover, though at higher costs and with longer processing times.

Our approach shows strong performance compared to existing automated clinical trial screening methods, although making direct comparisons with current literature is difficult due to the use of different datasets. Previous studies have explored rule-based techniques and supervised machine learning to organize trial eligibility criteria or align clinical trials with structured patient data. For example, Chen et al. developed an automated tool that matched patient biomarkers to clinical trials with high sensitivity and specificity. However, their model required manual extraction of data from unstructured text [10]. Similarly, Ni et al. created a logic/NLP-based pre-screening system for pediatric oncology patients that achieved over 90% sensitivity and specificity. However, their approach still relied on an advanced NLP pipeline and structured data entry [11].

Recently, there has been growing interest in using more advanced LLMs like ChatGPT to evaluate unstructured text, but these have primarily been tested on open-source datasets [19–21, 24]. Our EG approach stands out because it can be easily integrated into prompts by any clinician without requiring programming knowledge, and it can be written in everyday language. Moreover, our fully automated Self-Discover method performed just as well, eliminating the need for manual input. Our approach is versatile, as we previously demonstrated that it outperforms other methods explored in a clinical trial screening challenge [13, 14]. In our analysis, structured output with EG consistently yielded the best results for GPT-3.5, particularly in accuracy and sensitivity. However, more advanced approaches like Self-Discover, even with Custom Modules, were less effective for GPT-3.5, suggesting that this model benefits more from structured guidance. On the other hand, GPT-4 performed robustly across all prompting methods, with Self-Discover achieving the highest Youden Index. This indicates that GPT-4’s enhanced reasoning abilities make it well-suited for fully autonomous tasks. Nevertheless, GPT-4’s strong performance comes with higher computational costs and longer processing times, highlighting the trade-offs between efficiency and effectiveness.

Despite the promising performance for individual criteria, GPT was unable to identify patients who met all eligibility criteria when strict definitions were used. This is unsurprising given the large number of criteria used; even a model with 95% accuracy on each criterion by chance alone would miss nearly half of eligible cases (1-0.9514). Our alternative approach utilized GPT to identify the proportion of trial criteria met. This “proportional eligibility” approach was much more effective, achieving accuracy ranging from 0.67 to 0.80. In practice, this approach can be used to identify patients at the highest likelihood of being eligible for the trial who can then undergo manual chart review for pre-screening, increasing the efficiency of study coordinators who can begin chart reviews for those most likely to be eligible.

A significant advantage of our approach is the reduction in screening time compared to manual methods, which can take 30 to 45 minutes per patient [9, 12, 25]. Using LLMs, our prompting frameworks achieve fully automated screening in 1-3 minutes with GPT-3.5 and 8-12 minutes with GPT-4. This substantial reduction in screening time can greatly alleviate staff workload and costs, especially when EG is not required. The cost of screening a single patient ranged from $0.02 to $0.03 with GPT-3.5 and from $0.15 to $0.27 with GPT-4, making it a cost-effective solution that can focus high-cost, human efforts towards patients more likely to be eligible for a trial. While GPT-4 is more expensive per screening, its improved performance can justify the higher cost, particularly in highthroughput settings. Our costs appear similar ($1.55 per patient using GPT-4) to the only other study that reported such information, although it is difficult to make a direct comparison as there are likely different contributing factors [20].

Despite the advantages, an LLM-powered approach to clinical trial screening has some limitations compared to manual screening and other automated methods. While the cost of using LLMs can be significant due to charges per token by services like Microsoft Azure’s OpenAI, it remains much lower than the estimated $240 to $340 per-enrollment cost of manual screening [6, 26]. Rule-based and traditional NLP screening methods do not incur the same per-patient costs as our LLM-based approaches. Additionally, structured output and CoT prompting formats require a small training set and domain expertise to create EG, which can present a barrier to use. In contrast, Self-Discover methods do not have this limitation and can be implemented without domain expertise. Furthermore, hallucinations are a known concern for LLMs, requiring ongoing human review for confirmation of accuracy.

## 5 Conclusion

LLMs can be used to identify specific clinical trial criteria but have difficulties identifying patients who meet all criteria. Instead, using the proportion of criteria met to flag candidates for manual review is a viable approach. LLM performance varies by prompt, with GPT-4 generally outperforming GPT-3.5, but at higher costs and longer processing times. LLMs should complement, not replace, manual chart reviews for matching patients to clinical trials.

## Data Availability

All data produced in the present work are contained in the manuscript

## 6 Supplement A

### 6.1 Full Clinical Trial Criteria

**Inclusion Criteria** Inclusion criteria will be the same for Phase I and Phase II.

1. Pathologically proven diagnosis of stage I-IVB squamous cell carcinoma of the oral cavity, oropharynx, hypopharynx, or larynx status post gross total resection with pathology showing one or more of the following intermediate risk factors:

- T3/4 disease (AJCC 8th edition), positive lymph node(s), close margin(s), perineural invasion, and/or lymphovascular invasion
- Close margin(s) defined as either:

– Final patient margin of *<*5 mm without disease on ink OR
– Initial positive margin in the specimen regardless of the final patient margin (e.g. if resection margin on the initial specimen is positive, final patient margin after subsequent resections can be *≥*5 mm and still be considered close margin)
2. Age *≥*18 years
3. ECOG performance status 0-2
4. Women of child-bearing potential and men must agree to use adequate contraception (hormonal or barrier method of birth control; abstinence) prior to study entry, for the duration of study participation, and for 90 days following completion of therapy.
  - Medically acceptable birth control (contraceptives) includes:

(a) approved hormonal contraceptives (such as birth control pills, patch or ring; Depo-Provera, Implanon), or
(b) barrier methods (such as a condom or diaphragm) used with a spermicide
  - Should a woman become pregnant or suspect she is pregnant while participating in this study, she should inform her treating physician immediately.
  - A female of child-bearing potential is any woman (regardless of sexual orientation, having undergone a tubal ligation, or remaining celibate by choice) who meets the following criteria:

– Has not undergone a hysterectomy or bilateral oophorectomy; or
– Has not been naturally postmenopausal for at least 12 consecutive months (i.e., has had menses at any time in the preceding 12 consecutive months).
5. Negative serum or urine pregnancy test within 2 weeks before registration for women of childbearing potential.
6. Ability to understand and the willingness to sign a written informed consent.

#### Exclusion Criteria

1. Distant metastasis
2. Stage I and II (AJCC 8th edition) glottic squamous cell carcinoma
3. High risk factors following surgical resection requiring concurrent chemotherapy

a. Final positive margin(s) and/or extranodal extension
4. Feeding tube dependence (as defined in section 4.1.1) at baseline assessment
5. Synchronous non-skin cancer primaries outside of the oropharynx, oral cavity, larynx, and hypopharynx except for low- and intermediate-risk prostate cancer and synchronous well-differentiated thyroid cancer. For prostate cancer, patient should not be receiving active treatment. For thyroid cancer, thyroid surgery may occur before or after radiation treatment, provided all other eligibility criteria are met.
6. Prior invasive malignancy with an expected disease-free interval of less than 3 years
7. Prior radiotherapy to the region of the study cancer that would result in overlap of radiation fields
8. Subjects may not be receiving any other investigational agents for the treatment of the cancer under study.
9. Uncontrolled intercurrent illness including, but not limited to, ongoing or active infection, symptomatic congestive heart failure, unstable angina pectoris, cardiac arrhythmia, or psychiatric illness/social situations that, in the opinion of the investigator, would limit compliance with study requirements
10. Subjects must not be pregnant or nursing due to the potential for congenital abnormalities and the potential of this regimen to harm nursing infants.
11. History of severe immunosuppression, including HIV, and organ or autologous or allogeneic stem cell transplant

## 7 Prompts

### 7.1 General Prompt Template

**Role & Objective** You are a Clinical Trial Eligibility Screener. Your task is to review medical documents to assess patient eligibility for a specific clinical trial accurately. You can make accurate and definitive judgements based on medical records.

**Key Points Criteria:** *\*\*\*{criteria}\*\*\**

**Tips:** *###{tips}###*

#### Process

1. **Review Criteria:** Understand the trial criteria you are screening for (see ***) fully before starting.
2. **Examine Documents (see —):** Identify relevant sections based on the criteria. The documents provided comprise the entirety of relevant information from the patient’s EHR. Describe your thought process on what the evidence or lack of evidence means.
3. **Apply Tips:** Use provided tips (see ###) to aid your search. Apply judgment as needed.
4. **Assess Eligibility:** Determine if criteria are met or not met, based solely on document content (see —). When uncertain, determine the most likely answer and describe why.

Review the documents below and provide your answer.

## Documents

### 7.2 LLM Generated Guidance

#### CARCINOMA

**Requirement:** Patient must have a pathologically proven diagnosis of squamous cell carcinoma of the oral cavity, oropharynx, hypopharynx, or larynx status.

**Location:** Pathology Progress

#### Tips

- **Associated words for meeting the criterion:**

– biopsy, histopathology report, squamous cell carcinoma, oral cavity cancer, oropharyngeal cancer, hypopharyngeal cancer, laryngeal cancer, pathology confirmation, carcinoma diagnosis, SCC, malignant tumor, definitive diagnosis.
- **Associated words for not meeting the criterion**

– benign tumor, negative biopsy, no malignancy, non-cancerous, inconclusive pathology, different histology, adenocarcinoma, basal cell carcinoma, undiagnosed mass, no squamous cell carcinoma, absence of malignancy, non-squamous pathology.

### INTERMEDIATE-RISK

**Requirement:** Patient must have one or more intermediate-risk factors including: T3 or T4 disease per AJCC 8th edition staging, positive lymph node(s), close margin(s) as defined below, perineural invasion, and/or lymphovascular invasion. A ‘close margin’ is defined as less than 5 mm from the tumor edge without disease on ink, or any initial positive margin regardless of subsequent margin distance. Positive margins should not be considered close margins.

**Location:** Pathology

### Tips

- **Associated words for meeting the criterion:**

– T3 disease: “advanced primary tumor size,” “greater than 4 cm,” “lip or oral cavity involvement.”
– T4 disease: “very advanced primary tumor,” “invades adjacent structures,” “deep/extrinsic muscle of tongue,” “skin of face,” “maxillary sinus,” “bone erosion.”
– Positive lymph nodes: “nodal metastasis,” “regional lymph node involvement,” “N+,” “cervical lymphadenopathy.”
– Close margins: “resection margin ¡5 mm,” “tumor-free margin,” “narrow surgical margin,” “subsequent clear margin.”
– Perineural invasion: “nerve infiltration,” “neural spread,” “PNI positive.”
– Lymphovascular invasion: “vascular invasion,” “lymphatic spread,” “LVI positive.”
- **Associated words for not meeting the criterion:**
  – T1/T2 disease: “small primary tumor,” “less than 4 cm,” “limited to one site.”
  – Negative lymph nodes: “no nodal metastasis,” “N0,” “no regional lymph node involvement.”
  – Adequate margins: “resection margin *>*5 mm,” “clear margins,” “wide surgical margin.”
  – No perineural invasion: “PNI negative,” “no nerve infiltration.”
  – No lymphovascular invasion: “LVI negative,” “no vascular or lymphatic spread.”

## OVER-18

**Requirement:** Patient must have age greater than or equal to 18 years.

**Location:** Progress

### Tips

- **Associated words for meeting the criterion:**

– “adult,” “over 18,” “eligible age,” “legal age,” “mature,” “non-pediatric,” “age-qualified,” “consent-capable,” and “majority age.”
- **Words indicating the criterion is not met:**

– “minor,” “underage,” “below 18,” “ineligible age,” “child,” “adolescent,” “juvenile,” “not of age,” and “age-restricted.”

## ECOG

**Requirement:** Patient must have ECOG (Eastern Cooperative Oncology Group) performance status 0-2 or Karnofsky Performance Status (KPS) of 50-100.

**Location:** Progress

### Tips

- **Associated words for meeting the criterion:**

– fully active (ECOG 0), ambulatory (ECOG 1), capable of self-care (ECOG 2), moderate symptoms (KPS 50-70), requires assistance (KPS 50-60), able to care for most personal needs (KPS 70), limited in physically strenuous activity (KPS 80), normal activity with effort (KPS 90), and normal activity without restriction (KPS 100).
- **Words indicating the criterion is not met**:

– disabled (ECOG 3), limited self-care (ECOG 4), bedridden (ECOG 5), severely disabled (KPS below 50), hospitalization (KPS 30-40), and terminal illness (KPS 10-20).

## CONTRACEPTION

**Requirement:** Women of child-bearing potential and men must agree to use adequate contraception (hormonal or barrier method of birth control; abstinence) prior to study entry, for the duration of study participation, and for 90 days following completion of therapy.

**Location:** Progress

### Tips

- **Associated words for meeting the criterion:**

– consent, agreement, compliance, contraception, hormonal methods, birth control pills, IUD, barrier methods, condoms, diaphragm, abstinence, understanding, informed, duration, study participation, post-therapy, 90 days, reproductive age, fertility awareness, vasectomy, tubal ligation.
- **Associated words for not meeting the criterion:**

– non-compliance, refusal, disagreement, non-consent, unprotected sex, contraceptive failure, lack of understanding, non-adherence, withdrawal method, fertility, pregnancy risk, contraception contraindications, non-abstinence, postpartum.

## NEGATIVE-PREGNANCY

**Requirement:** Patient must have a negative serum or urine pregnancy test within 2 weeks before registration for women of childbearing potential.

**Location:** Labs

### Tips

- **Associated words for meeting the criterion:**

– negative pregnancy test, non-pregnant, serum test, urine test, within 2 weeks, pre-registration, eligibility confirmed, women of childbearing potential, exclusion of pregnancy, laboratory confirmation, test date, test result.
- **Associated words for not meeting the criterion:**

– positive pregnancy test, pregnant, ineligible, beyond 2 weeks, post-registration, women of childbearing potential, pregnancy confirmed, disqualification, outdated test, pending test result, test not performed.

## NO-DISTANT-METASTASIS

**Requirement:** Patient must not have distant metastasis outside of the head and neck region.

**Location:** Progress Pathology

### Tips

- **Associated words for meeting the criterion:**

– localized, confined, regional, non-metastatic, NED (no evidence of disease), clear scans, remission, within head and neck.
- **Associated words for not meeting the criterion:**

– metastatic disease, spread, secondary tumors, distant lesions, M1 (metastasis present), extracervical, systemic involvement, beyond head and neck, advanced disease.

## NO-GLOTTIC

**Requirement:** Patient must not have stage I or II (AJCC 8th edition) glottic squamous cell carcinoma.

**Location:** Pathology Progress

### Tips

- **Associated words for meeting the criterion:**

– Stage III, Stage IV, Stage IVB, advanced, non-glottic, supraglottic, subglottic, transglottic, oropharyngeal, oral cavity, hypopharyngeal, laryngeal, T3, T4, N+, extrinsic muscles of the larynx, vocal cord fixation, thyroid cartilage erosion, extensive tumor.
- **Associated words for not meeting the criterion:**

– Stage I, Stage II, early-stage, glottic, limited to vocal cords, vocal cord mobility intact, confined to larynx, no extralaryngeal spread, AJCC 8th edition, small tumor, minimal invasion.

## NO-HIGH-RISK

**Requirement:** Patient must not have high-risk factors following surgical resection including positive surgical margins or extracapsular extension of lymph node metastasis.

**Location:** Pathology

### Tips

- **Associated words for meeting the criterion:**

– negative margins, clear margins, encapsulated lymph nodes, no extracapsular spread, negative extracapsular extension, intact lymph node capsule.
- **Associated words for not meeting the criterion:**

– positive margins, involved margins, positive surgical margins, extracapsular extension, extracapsular spread, lymph node metastasis with extracapsular extension, breached lymph node capsule.

## NO-FEEDING-TUBE

**Requirement:** Patient must not have a feeding tube dependence at baseline assessment.

**Location:** Progress Img Narrative

### Tips

- **Associated words for meeting the criterion:**

– independent feeding, oral intake, self-feeding, no feeding tube, swallowing function intact, adequate nutrition orally, no enteral support.
- **Associated words for not meeting the criterion:**

– feeding tube present, gastrostomy, PEG (percutaneous endoscopic gastrostomy), enteral feeding, tube dependence, dysphagia requiring tube, nutritional support via tube, inability to swallow, NG tube (nasogastric tube).

## NO-SYNCHRONOUS

**Requirement:** Patient must not have synchronous non-skin cancer primaries outside of the oropharynx, oral cavity, larynx, and hypopharynx except for low- and intermediate-risk prostate cancer and synchronous well-differentiated thyroid cancer.

**Location:** Progress

### Tips

- **Associated words for meeting the criterion:**

– “single primary,” “exclusive,” “confined,” “localized,” “oral cavity,” “oropharynx,” “larynx,” “hypopharynx,” “low-risk prostate cancer,” “intermediate-risk prostate cancer,” “well-differentiated thyroid cancer,” “synchronous.”
- **Associated words for not meeting the criterion:**

– “multiple primaries,” “metastatic,” “disseminated,” “additional cancer,” “high-risk prostate cancer,” “poorly-differentiated thyroid cancer,” “advanced-stage,” “extraneous malignancy,” “breast cancer,” “colon cancer,” “lung cancer,” “synchronous non-skin primaries.”

## NO-SHORT-DISEASE-FREE

**Requirement:** Patient must not have prior invasive malignancy with an expected disease-free interval of less than 3 years.

**Location:** Progress

### Tips

- **Associated words for meeting the criterion:**

– disease-free, remission, non-recurrent, stable, no history of cancer, no other malignancies, clear medical history, cancer-free for 3+ years.
- **Associated words for not meeting the criterion:**

– recent cancer diagnosis, active malignancy, recurrence, metastasis, ongoing treatment, recent remission, history of cancer within 3 years, second primary cancer, short disease-free interval.

## NO-PRIOR-RADIO

**Requirement:** Patient must not have had prior radiotherapy or radiation to the head or neck.

**Location:** Progress

### Tips

- **Associated words for meeting the criterion:**

– radiotherapy-naive, radiation-naive, no previous radiation, no history of radiotherapy, radiation-free history, untreated by radiotherapy, no prior radiation treatment.
- **Associated words for not meeting the criterion:**

– previously irradiated, history of radiotherapy, prior radiation treatment, postradiotherapy, prior head and neck radiation, radiation-treated, prior therapeutic radiation.

## NO-OTHER-AGENTS

**Requirement:** Patient must not be receiving any other investigational agents for the treatment of the cancer under study.

**Location:** Progress

### Tips

- **Associated words for meeting the criterion:**

– “not enrolled,” “no concurrent trials,” “exclusive participation,” “single study involvement,” “no investigational drugs,” “no other treatments,” “sole therapy,” “monotherapy.”
- **Associated words for not meeting the criterion:**

– “concurrent enrollment,” “participating in other trials,” “receiving other investigational agents,” “multiple study involvement,” “combination therapy,” “additional investigational drugs,” “polytherapy.”

## NO-INCURRENT-ILLNESS

**Requirement:** Patient must not have uncontrolled intercurrent illness including, but not limited to, ongoing or active infection, symptomatic congestive heart failure, unstable angina pectoris, cardiac arrhythmia, psychiatric illness/social situations that would limit compliance with study requirements.

**Location:** Progress

### Tips

- **Associated words for meeting the criterion:**

– Stable
– Controlled
– Asymptomatic
– Euthymic
– Compliant
– Managed chronic conditions
– No active infection
– Stable cardiac status
– Adequate social support
- **Associated words for not meeting the criterion:**

– Uncontrolled diabetes
– Active infection
– Acute exacerbation
– Decompensated heart failure
– Unstable angina
– Severe arrhythmia
– Psychiatric crisis
– Non-compliance
– Social instability
– Unmanaged hypertension

## NO-PREGNANCY

**Requirement:** Patient must not be pregnant or nursing due to the potential for congenital abnormalities and the potential of this regimen to harm nursing infants.

**Location:** Progress

### Tips

- **Associated words for meeting the criterion:**

– not pregnant, non-pregnant, not nursing, not lactating, negative pregnancy test, contraception use, post-menopausal, vasectomized partner, abstinence.
- **Associated words for not meeting the criterion:**

– pregnant, positive pregnancy test, lactating, breastfeeding, nursing, potential mother, childbearing potential without contraception, pre-menopausal without contraception.

## NO-IMMUNOSUPPRESSION

**Requirement:** Patient must not have a history of severe immunosuppression, including HIV, and organ or autologous or allogeneic stem cell transplant.

**Location:** Progress

### Tips

- **Associated words for meeting the criterion:**

– Immunocompetent
– Negative HIV status
– No transplant history
– Healthy immune system
– Non-immunosuppressed
- **Associated words for not meeting the criterion:**

– HIV positive
– Immunosuppressed
– Organ transplant recipient
– Stem cell transplant (autologous or allogeneic)
– Immune deficiency
– Chronic immunosuppressive therapy
– Severe immunodeficiency

### 7.3 Expert Guidance Example

#### CARCINOMA

**Location:** Pathology Progress

#### Tips

- Focus on detailed anatomical subsites within the oral cavity (lip, oral tongue, floor of mouth, buccal mucosa, upper and lower gum, retromolar trigone, and hard palate), oropharynx (base of the tongue to the epiglottis, tonsils, pharyngeal walls, and soft palate), larynx (thyroid cartilage, cricoid cartilage, total larynx, vocal cords, supraglottic, subglottic, glottic larynx, arytenoids, aryepiglottic folds, vestibular folds, and glossoepiglottic fold), and hypopharynx (piriform sinuses, lateral and posterior pharyngeal walls, and posterior surfaces of the larynx). If no evidence of squamous cell carcinoma of the oral cavity, oropharynx, hypopharynx, or larynx is found, then the criteria is not met.

#### INTERMEDIATE-RISK

**Requirement:** Patient must have one or more Intermediate-Risk Factors including: T3 or T4 disease per AJCC 8th edition staging, positive lymph node(s), close margin(s) as defined below, perineural invasion, and/or lymphovascular invasion. A ‘close margin’ is defined as less than 5 mm from the tumor edge without disease on ink, or any initial positive margin regardless of subsequent margin distance. Positive margins should not be considered close margins.

**Location:** Pathology

#### Tips

- Look to pathology reports from resections for information on margin status, and pathology reports from biopsies for information on perineural invasion, lymphovascular invasion, and lymph nodes. Clinic notes may supplement missing details. Key staging specifics:

– Oropharynx: T3 *>* 4 cm or extending to the epiglottis; T4a invades adjacent structures; T4b involves critical areas like the base of the skull.
– Oral Cavity: T3 *>* 4 cm or with significant depth; T4a invades facial bones/deep structures; T4b spreads to skull base.
– Larynx: T3 involves vocal cord paralysis; T4a spreads to nearby tissues; T4b extends to prevertebral space or encases arteries.
– Hypopharynx: T3 *>* 4 cm or extends to larynx/esophagus; T4a invades nearby structures; T4b involves prevertebral fascia or chest.
- You do not need all of the above information to rule a patient as meeting the criteria. If sufficient evidence exists to suggest that the criteria is met, then the criteria is met. If no evidence of intermediate risk factors are found, then the criteria is not met.

#### OVER-18

**Requirement:** Patient must have age greater than or equal to 18 years.

**Location:** Progress

#### Tips

- Look for the patient’s age that may be explicitly stated in the clinic note. Alternatively, evaluate the date of birth (DOB); the year should be before 2006.

#### ECOG

**Location:** Progress

#### Tips

- When assessing performance status, specifically search for mentions of ‘ECOG,’ ‘Karnofsky Performance Status,’ ‘KPS,’ or the full terms spelled out within the clinic consultation notes, particularly in the physical exam section. These indicators are key to determining the patient’s functional status accurately.
- If not directly stated, infer the status from the patient’s activity level descriptions or mentions of physical abilities (walking, standing, independence). For instance:

– Able to walk/ambulate without assistance suggests ECOG 0 or 1.
– Bedridden indicates a performance status likely at ECOG 3 or 4.
- Example:
  – Text: Physical Examination: See vitals in epic Wt Readings from Last 3 Encounters: 02/02/24 113 lb 9.6 oz (51.5 kg) 08/21/23 122 lb 6.4 oz (55.5 kg) 08/09/23 120 lb (54.4 kg) ECOG: 2 GENERAL: NAD HEAD/NECK: Notable mass extending across right side of face and neck LUNGS: Breathing comfortably HEART: Adequate perfusion NEUROLOGIC: AOx3, speech fluent, hearing intact to conversational tone, moves all extremities. CNII-XII grossly intact. Normal gait. EXTREMITIES: There is no upper or lower extremity edema. Assessment: The ECOG performance status is 2, which meets the eligibility criteria for this trial. The patient’s ability to walk without assistance and the absence of bedridden status align with the ECOG 0-2 requirement.

#### CONTRACEPTION

**Location:** Progress

#### Tips

- Medically acceptable birth control (contraceptives) includes:

– 1) approved hormonal contraceptives (such as birth control pills, patch or ring; Depo-Provera, Implanon), or
– 2) barrier methods (such as a condom or diaphragm) used with a spermicide.
- Criteria is also met if the patient is a woman but has undergone a hysterectomy or bilateral oophorectomy or has been naturally postmenopausal for at least 12 consecutive months. If the patient is a male, criteria is also met if the patient has undergone a successful vasectomy however if nothing is mentioned then can assume criteria is met (men only). If nothing explicitly contradicts this criteria, then this criteria is met.

#### NEGATIVE-PREGNANCY

**Location:** Labs

#### Tips

- If the patient is a male, then this criteria is not applicable but can put true/met. Otherwise, look for a pregnancy test result in the lab section or in the lab section of the clinic note.

#### NO-DISTANT-METASTASIS

**Requirement:** Patient must not have distant metastasis outside of the head and neck region.

**Location:** Progress Pathology

#### Tips

- Review clinical notes and imaging reports for AJCC staging and specific mentions of distant metastasis. Stages I-IVA generally indicate eligibility, as they typically do not involve distant metastasis. Any mention of ‘Stage IVB’ or ‘M1’ (which explicitly indicates distant metastasis) means the criteria are not met. Focus on identifying any descriptions of locoregional disease without signs of spread to distant organs like the lungs, liver, or bones. Confirmation of no distant metastasis (N0, M0) aligns with the criterion. If there is cancer in the head or neck without other distant metastatic disease, then the criteria is met. If there is no mention of any metastases, then the criterion is met.
- Additionally, when interpreting PET/CT scans, focus on the ‘Impression’ and ‘Findings’ sections for indications of FDG avid lesions. Ensure there are no signs of hypermetabolic activity suggesting distant metastasis outside the head and neck, such as in the lungs, bones, abdomen, liver, or pelvis. The presence of hypermetabolic lymph nodes within the neck is acceptable, but the key is to exclude FDG avidity in distant organs or tissues.

#### NO-GLOTTIC

**Location:** Pathology Progress

#### Tips

- Identify glottic cancer beyond early stages by excluding Tis (top layer only), T1 (within vocal cords, with T1a affecting one and T1b affecting both vocal cords, both allowing normal movement), and T2 (extends to adjacent areas with possible movement impairment). Evidence of more extensive disease or involvement in the oral cavity, oropharynx, hypopharynx, or larynx without Stage I or II classification suggests later-stage cancer. Focus on confirming disease scope beyond the vocal cords for accurate assessment.
- Having T1-T2 disease means the criteria are not met, as these stages are considered early-stage glottic cancer. Conversely, having T3, T4, N1, Stage III or IV disease indicates the criteria are met, as these stages signify the cancer is beyond the early stages. If no mention of glottic cancer is found, then this criteria is met. If evidence of cancer is found but it is not explicitly described as glottic, then the criteria is met. If glottic cancer is mentioned but no stage is explicitly stated, infer the stage from the disease description.

#### NO-HIGH-RISK

**Location:** Pathology

#### Tips

- Presence of high-risk factors, specifically positive surgical margins or extracapsular extension in lymph node metastasis, means the criteria are not met. If initial surgery had positive margins but a subsequent resection yields negative margins, the criteria are met. Conversely, if a repeat resection still shows positive margins, the criteria remain unmet. Assess pathology reports and clinic notes for any mentions of these high-risk factors to accurately determine eligibility. If no evidence of high risk factors are found, then the criteria is met.

#### NO-FEEDING-TUBE

**Requirement:** Patient must not have a feeding tube dependence at baseline assessment.

**Location:** Progress Img Narrative

#### Tips

- Review clinic notes carefully for any explicit mention of feeding tube use. If there is documentation of a feeding tube, the patient does not meet this criterion. In cases where clinic notes do not mention a feeding tube, it’s safe to assume the patient is not dependent on one, and thus, meets the criterion. Additionally, consider checking the patient’s nutritional assessment or treatment plan sections within the notes for comprehensive evaluation.
- Additionally, it’s important to review imaging reports for any indications of feeding tube presence, as these reports may incidentally note feeding tubes not mentioned in clinic notes. If no mention of feeding tube, then this criteria is met.

#### NO-SYNCHRONOUS

**Location:** Progress

#### Tips

- Review clinic notes and reports for any active synchronous cancers, which are two or more primary cancers occurring at the same time in different locations. Active cancers outside the oropharynx, oral cavity, larynx, and hypopharynx are not permissible. When reviewing clinic notes and reports, specifically identify any active instances of lung, gastrointestinal, brain, bladder, ovarian, cervical cancers, sarcoma, or lymphoma, as these are not permissible for eligibility. If these cancers are mentioned as historical and currently inactive, the criteria are considered met. If no mention of the non-permissible cancer then criteria is also met. Again patients are allowed to have low/intermediate risk prostate cancer or well differentiated thyroid cancer.

#### NO-SHORT-DISEASE-FREE

**Location:** Progress

#### Tips

- Review patient history for any invasive malignancies within the last three years, focusing on aggressive forms like lung, pancreatic, breast, colorectal, melanoma, glioblastoma, and any metastatic cancers. These malignancies typically have shorter disease-free intervals. Exclude prostate cancer, well-differentiated thyroid cancer, and squamous cell skin cancer from this assessment. Look for mentions of recurrence, as well as mentions of any cancer primaries. The record dates should be compared, and if there is a recurrence less than 3 years after the mention of a cancer primary, then the criterion is not met. If there’s no mention of such cancers, or only the excluded types are present, then the criteria are considered met.

#### NO-PRIOR-RADIO

**Requirement:** Patient must not have had prior radiotherapy or radiation to the head or neck.

**Location:** Progress

#### Tips

- Examine patient records for mentions of prior radiation specifically targeting the head or neck region, as noted in clinic or radiation oncology documentation. Radiation treatments to body areas outside the head and neck, like the brain, thorax, chest, lungs, prostate, liver, or pelvis, are not considered a contraindication for this criterion. The focus is on identifying any radiation therapy that overlaps with the cancer study region. If there is mention past ‘treatment’ to the head or neck in conjunction with a dosage measured in Gy, then the criterion is not met. Ignore any instances of radiotherapy being planned or consented to. Only consider cases where a patient is documented as receiving radiotherapy in the past. If no mention of radiation or radiotherapy is found, the criteria is met.

#### NO-OTHER-AGENTS

**Location:** Progress

#### Tips

- Specifically, look in the clinic notes if the patient is currently enrolled in another study for the management of their head and neck cancer. If they are enrolled in a clinical trial for head and neck cancer, then this criteria is not met. If no mention of other studies or investigation agents are found, the criteria is met.

#### NO-INCURRENT-ILLNESS

**Location:** Progress

#### Tips

- If the patient has a history of these conditions but they are not active, then this criteria is met. If the patient has active history but is properly controlled with medication, then this criteria is met. If no mention of issues with compliance then criteria is met. Look for mention of these conditions in the clinic note. If no mention of these conditions are found, the criteria is met.

#### NO-PREGNANCY

**Location:** Progress

#### Tips

- If the patient is a woman then evaluate clinic notes and lab results that would suggest pregnancy or breastfeeding. If the patient is male then this criteria is not applicable but can put true/met. Unless there is mention of pregnancy or breastfeeding, then this criteria is met.

#### NO-IMMUNOSUPPRESSION

**Requirement:** Patient must not have a history of severe immunosuppression, including HIV, and organ or autologous or allogeneic stem cell transplant.

**Location:** Progress

#### Tips

- Look for mention of these conditions in the clinic note. If patient has a history of HIV specifically but it is well controlled with medication then this criteria is met. If the charts do not mention these conditions, then this criteria is met.

### 7.4 Self-Discover Prompts and Modules

#### Chain of Prompts

Select Prompt:

- In order to solve the given task:

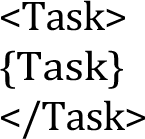 Select up to 5 modules that are crucial for solving the tasks above from all the reasoning module descriptions given below:

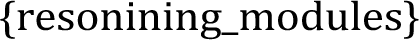

#### Adapt Prompt

- Rephrase and specify each reasoning module so that it better helps solving the task:

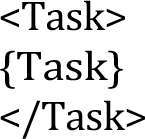 Adapt each reasoning module description to better solve the task:

#### Implement Prompt

- Without working out the full solution, create an actionable reasoning structure for the task using these adapted reasoning modules:

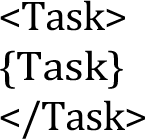

#### Final Prompt

- Using the following reasoning structure, solve the given task, providing your final answer:

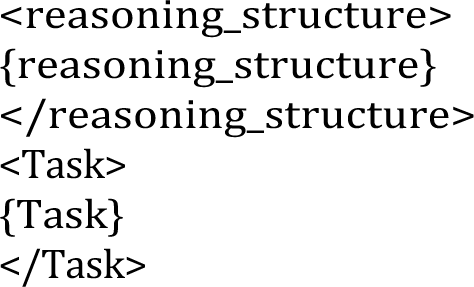 Do not simply repeat the prompts. Provide your reasoning for each step and your final answer below:

#### Reasoning Mo dules

1. How could I devise an experiment to help solve that problem?
2. Make a list of ideas for solving this problem, and apply them one by one to the problem to see if any progress can be made.
3. How could I measure progress on this problem?
4. How can I simplify the problem so that it is easier to solve?
5. What are the key assumptions underlying this problem?
6. What are the potential risks and drawbacks of each solution?
7. What are the alternative perspectives or viewpoints on this problem?
8. What are the long-term implications of this problem and its solutions?
9. How can I break down this problem into smaller, more manageable parts?
10. Critical Thinking: This style involves analyzing the problem from different perspectives, questioning assumptions, and evaluating the evidence or information available. It focuses on logical reasoning, evidence-based decision-making, and identifying potential biases or flaws in thinking.
11. Try creative thinking, generate innovative and out-of-the-box ideas to solve the problem. Explore unconventional solutions, thinking beyond traditional boundaries, and encouraging imagination and originality.
12. Seek input and collaboration from others to solve the problem. Emphasize teamwork, open communication, and leveraging the diverse perspectives and expertise of a group to come up with effective solutions.
13. Use systems thinking: Consider the problem as part of a larger system and understanding the interconnectedness of various elements. Focus on identifying the underlying causes, feedback loops,and interdependencies that influence the problem, and developing holistic solutions that address the system as a whole.
14. Use Risk Analysis: Evaluate potential risks, uncertainties, and tradeoffs associated with different solutions or approaches to a problem. Emphasize assessing the potential consequences and likelihood of success or failure, and making informed decisions based on a balanced analysis of risks and benefits.
15. Use Reflective Thinking: Step back from the problem, take the time for introspection and self-reflection. Examine personal biases, assumptions, and mental models that may influence problem-solving, and being open to learning from past experiences to improve future approaches.
16. What is the core issue or problem that needs to be addressed?
17. What are the underlying causes or factors contributing to the problem?
18. Are there any potential solutions or strategies that have been tried before? If yes, what were the outcomes and lessons learned?
19. What are the potential obstacles or challenges that might arise in solving this problem?
20. Are there any relevant data or information that can provide insights into the problem? If yes, what data sources are available, and how can they be analyzed?
21. Are there any stakeholders or individuals who are directly affected by the problem? What are their perspectives and needs?
22. What resources (financial, human, technological, etc.) are needed to tackle the problem effectively?
23. How can progress or success in solving the problem be measured or evaluated?
24. What indicators or metrics can be used?
25. Is the problem a technical or practical one that requires a specific expertise or skill set? Or is it more of a conceptual or theoretical problem?
26. Does the problem involve a physical constraint, such as limited resources, infrastructure, or space?
27. Is the problem related to human behavior, such as a social, cultural, or psychological issue?
28. Does the problem involve decision-making or planning, where choices need to be made under uncertainty or with competing objectives?
29. Is the problem an analytical one that requires data analysis, modeling, or optimization techniques?
30. Is the problem a design challenge that requires creative solutions and innovation?
31. Does the problem require addressing systemic or structural issues rather than just individual instances?
32. Is the problem time-sensitive or urgent, requiring immediate attention and action?
33. What kinds of solution typically are produced for this kind of problem specification?
34. Given the problem specification and the current best solution, have a guess about other possible solutions.
35. Let’s imagine the current best solution is totally wrong, what other ways are there to think about the problem specification?
36. What is the best way to modify this current best solution, given what you know about these kinds of problem specification?
37. Ignoring the current best solution, create an entirely new solution to the problem.
38. Let’s think step by step.
39. Let’s make a step by step plan and implement it with good notion and explanation.

### Custom Reasoning Modules

1. Gather and analyze relevant medical information from various sources (e.g., medical notes, lab values, pathology reports, imaging studies) to form a comprehensive understanding of the patient’s condition.
2. Identify the key clinical questions or problems that need to be addressed based on the available information.
3. Use critical thinking to analyze the clinical problem from different perspectives, question assumptions, and evaluate the available evidence.
4. Consider how the current clinical problem fits within the larger context of the patient’s overall health and medical history (systems thinking).
5. Simplify complex clinical problems by breaking them down into smaller, more manageable components.
6. Identify the key assumptions underlying the clinical problem and assess their validity based on the available evidence.
7. Generate a list of potential diagnoses or treatment options based on the clinical information and current medical knowledge.
8. Analyze the potential risks, benefits, and drawbacks of each diagnostic or treatment option (risk analysis).
9. Consider alternative perspectives or viewpoints on the clinical problem, such as those of other healthcare professionals, the patient, or their family members.
10. Use creative thinking to generate innovative or unconventional diagnostic or treatment approaches, when appropriate.
11. Critically evaluate the current working diagnosis or treatment plan and consider alternative explanations or approaches.
12. Use evidence-based medicine principles to select the most appropriate diagnostic tests or treatments based on the best available research evidence, clinical expertise, and patient preferences.
13. Develop a step-by-step diagnostic or treatment plan with clear reasoning and justification for each decision point.
14. Communicate the clinical reasoning process and decision-making to other healthcare team members, the patient, and their family in a clear and understandable manner.
15. Monitor the patient’s response to diagnostic tests or treatments and adjust the plan as needed based on their clinical course and any new information that becomes available.
16. Continuously reassess the clinical problem and treatment plan in light of new data, research findings, or changes in the patient’s condition.
17. Recognize and address any potential biases or flaws in the clinical reasoning process, such as anchoring bias or premature closure.
18. Reflect on patient outcomes and use them to inform future clinical decision-making and practice improvement.
19. Engage in lifelong learning to stay current with the latest medical knowledge and advances in diagnostic and treatment approaches.
20. Collaborate with other healthcare professionals to gather diverse perspectives and expertise in solving complex clinical problems.

### Text-Specific Modules

#### Pathology Reports

1. Contextualize the findings: Consider the patient’s clinical history and reason for the procedure to provide context for interpreting the pathology report.
2. Analyze gross and microscopic descriptions: Carefully examine the gross and microscopic descriptions, noting any abnormalities or key features that may guide diagnosis.
3. Interpret diagnostic summary: Focus on the diagnostic summary or conclusion, which provides the most critical information for clinical decision-making.
4. Integrate with other findings: Correlate the pathological findings with clinical presentation and other diagnostic tests to form a comprehensive understanding of the patient’s condition.
5. Consider additional studies: Identify any additional studies that may refine the diagnosis or provide prognostic information, when appropriate.

#### Physician Clinical Notes

1. Identify the main concern: Clearly define the patient’s chief complaint and reason for seeking medical attention.
2. Assess relevant background: Review pertinent past medical history, medications, allergies, and family history to inform the current clinical problem.
3. Analyze physical examination: Carefully consider the physical examination findings, particularly any abnormalities or signs related to the presenting problem.
4. Evaluate clinical impression: Assess the physician’s differential diagnosis or clinical impression based on the available information, and consider alternative explanations.
5. Interpret plan and follow-up: Analyze the physician’s assessment and plan, including any diagnostic tests, treatments, or referrals, and ensure appropriate follow-up.

#### Radiology Reports

1. Consider clinical context: Review the patient’s clinical history and indication for the imaging study to guide interpretation of the findings.
2. Systematically assess images: Methodically analyze the imaging findings, focusing on any abnormalities, lesions, or suspicious areas that may be relevant to the clinical question.
3. Interpret radiologist’s impression: Carefully consider the radiologist’s impression or conclusion, which summarizes the main findings and their potential significance.
4. Correlate with other data: Integrate the imaging findings with the patient’s clinical presentation, physical examination, and other diagnostic tests to refine the differential diagnosis.
5. Plan further evaluation: Determine if any additional imaging or procedures are recommended based on the initial findings, and collaborate with the treating physician to plan next steps.

#### Lab Reports

1. Identify test purpose: Clearly define the reason for ordering each lab test and its role in the diagnostic workup.
2. Compare to reference ranges: Systematically compare the patient’s lab values to the appropriate reference ranges, highlighting any abnormalities or critical values.
3. Interpret in clinical context: Analyze the lab results in the context of the patient’s clinical presentation, considering potential confounding factors and limitations of the tests.
4. Recognize patterns and implications: Identify any patterns or constellations of lab abnormalities that may suggest specific diagnoses or organ dysfunction, and consider their implications for patient management.
5. Monitor trends over time: Assess trends in the patient’s lab values over time to guide decisions about disease progression, treatment response, and potential complications.

## Supplement C

**Table C1.** Distribution of “met” and “not met” labels among patients who were ineligible.

| **Criterion** | **Met** | **Not Met** |
| --- | --- | --- |
| Carcinoma | 10 (34%) | 19 (66%) |
| Intermediate Risk Factors | 13 (45%) | 16 (55%) |
| ECOG $\leq 2$ | 28 (97%) | 1 (3%) |
| No Distant Metastasis | 26 (90%) | 3 (10%) |
| No Glottic Cancer | 27 (93%) | 3 (7%) |
| No High Risk Factors | 27 (93%) | 3 (7%) |
| No Synchronous Cancer | 28 (97%) | 1 (3%) |
| No Short Disease-Free Interval | 24 (83%) | 5 (17%) |
| No Prior Radiotherapy | 26 (90%) | 3 (10%) |
| No Investigational Agents | 29 (100%) | 0 (0%) |
| No Intercurrent Illness | 27 (93%) | 3 (7%) |
| No Pregnancy | 29 (100%) | 0 (0%) |
| No Immunosuppression | 28 (97%) | 1 (3%) |
| No Feeding Tube Dependence | 28 (97%) | 1 (3%) |

**Table C2:**
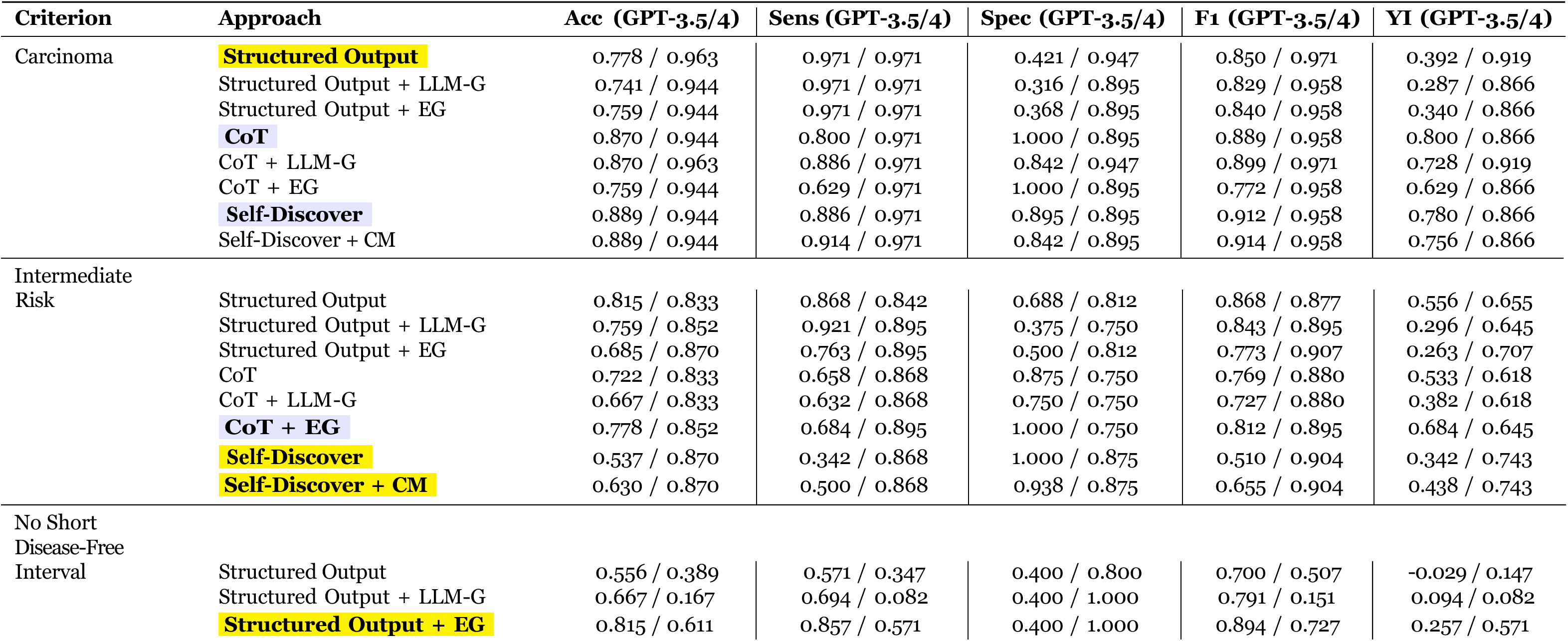
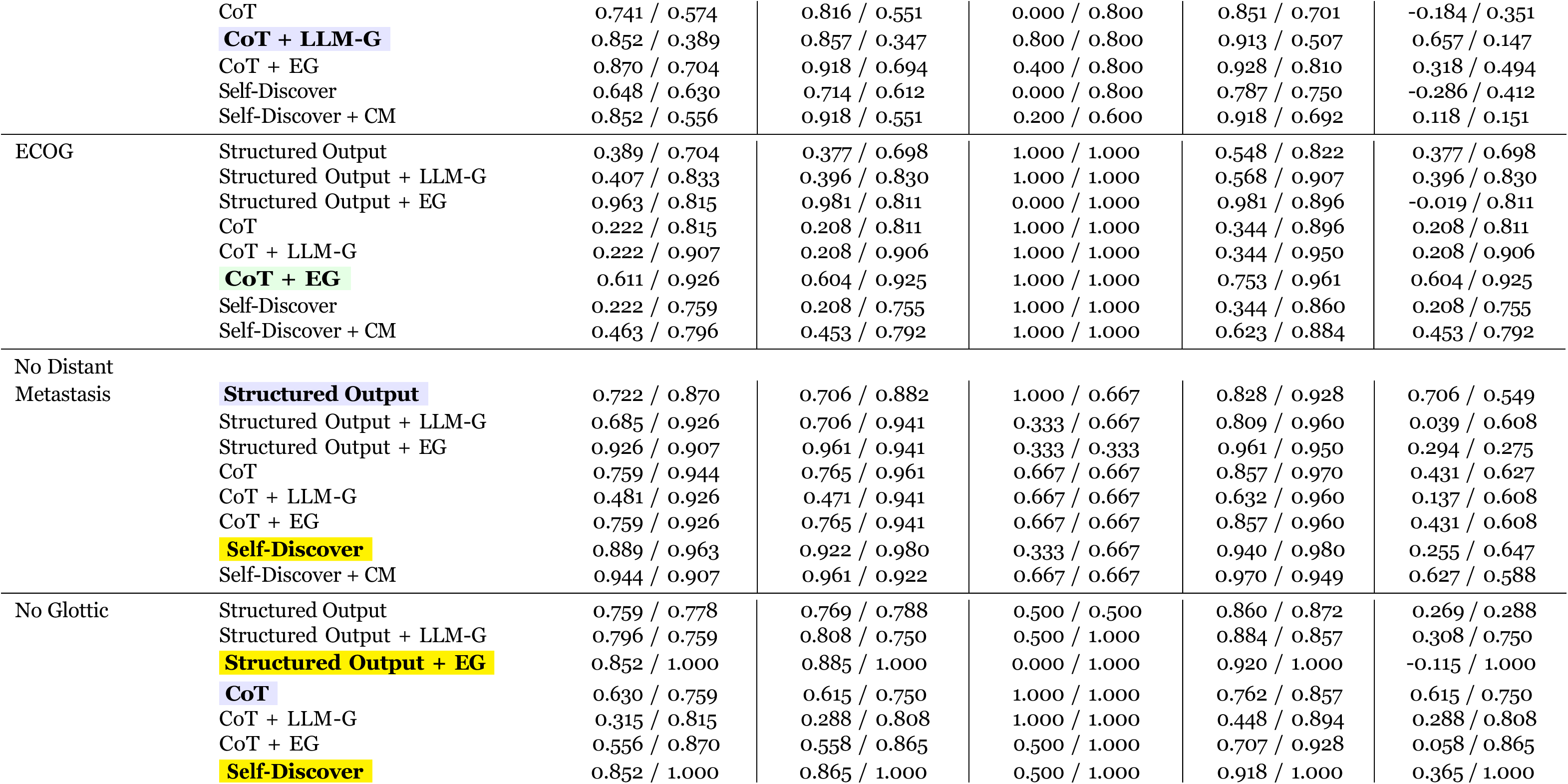
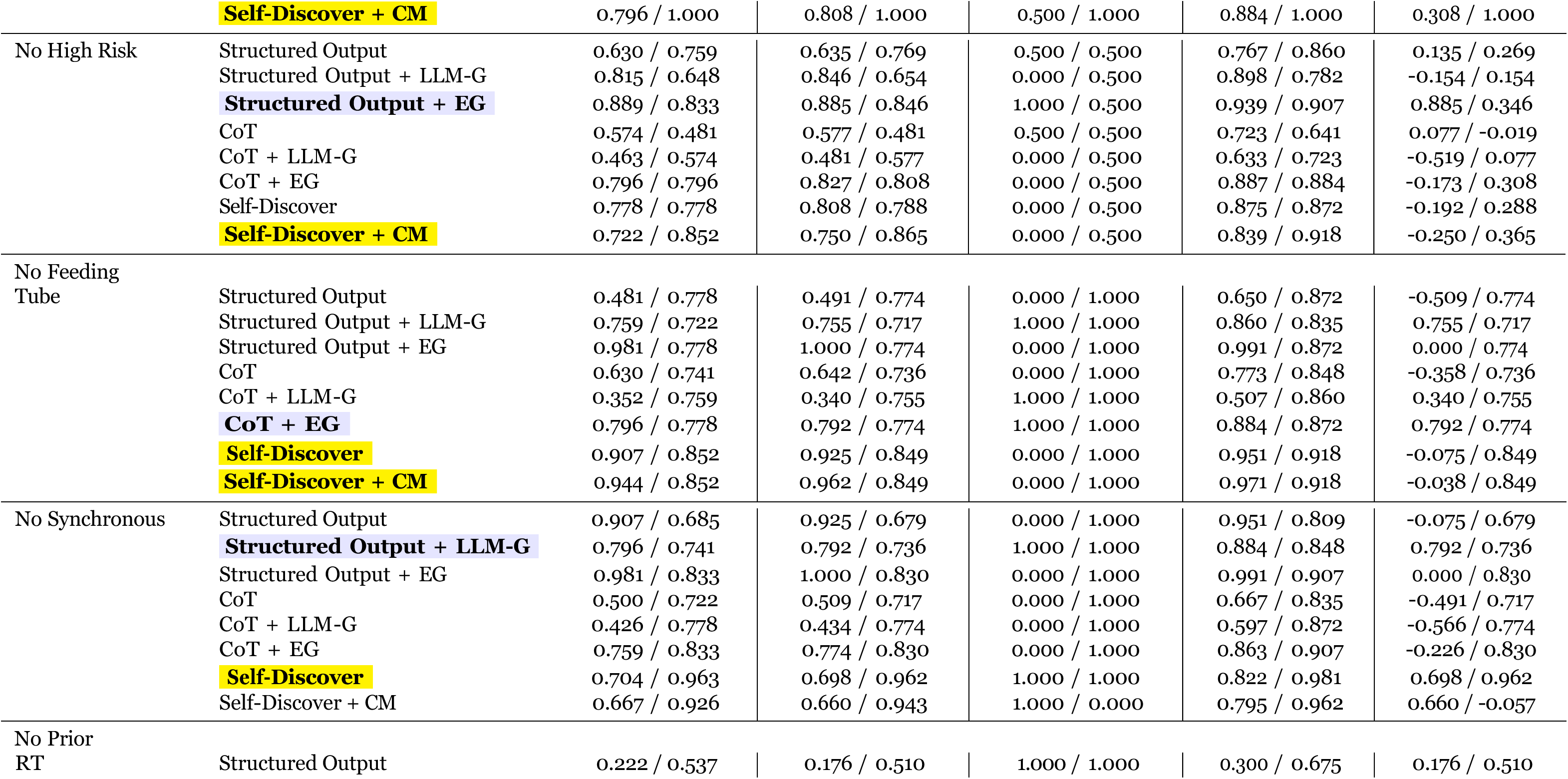
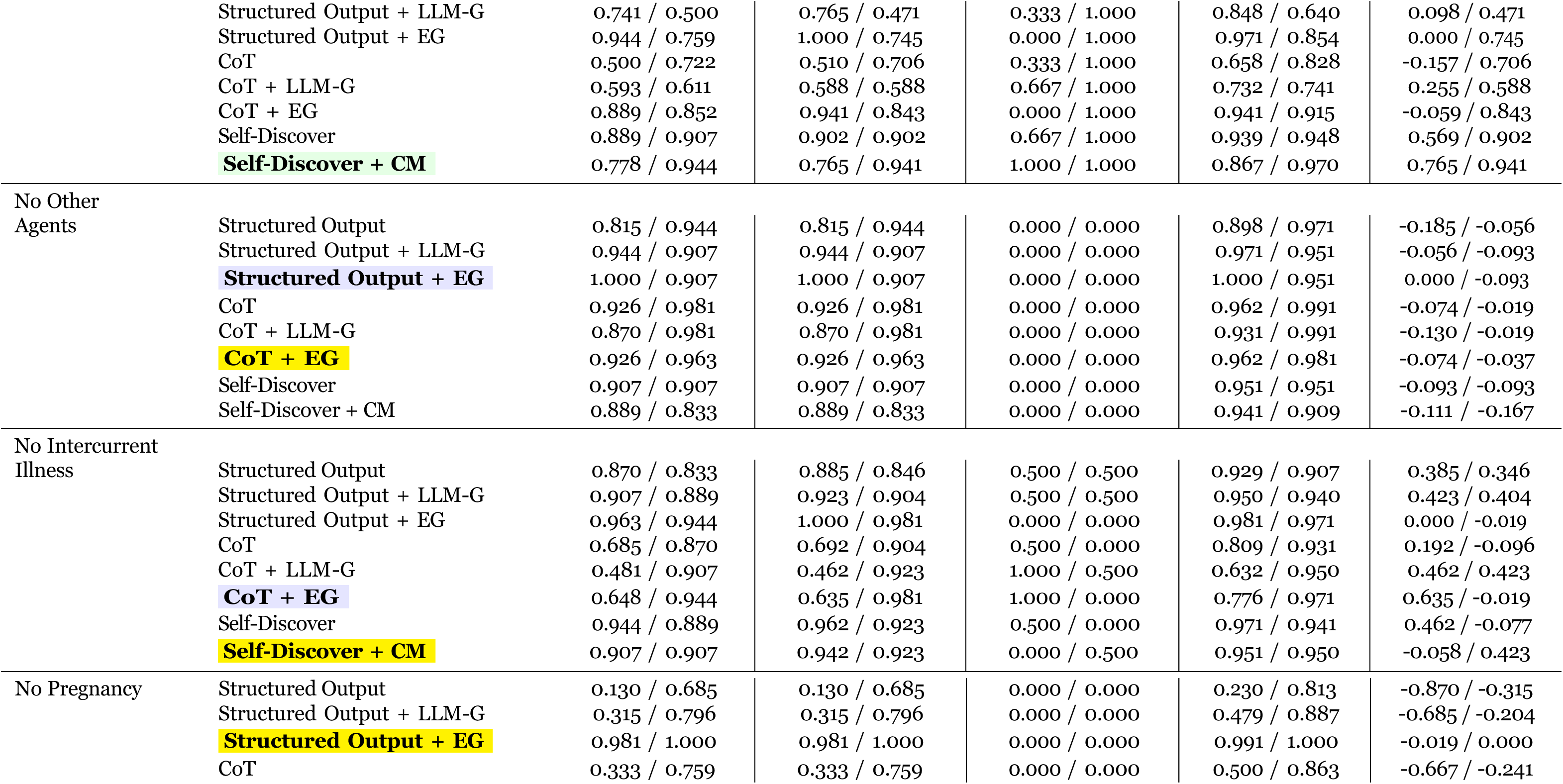
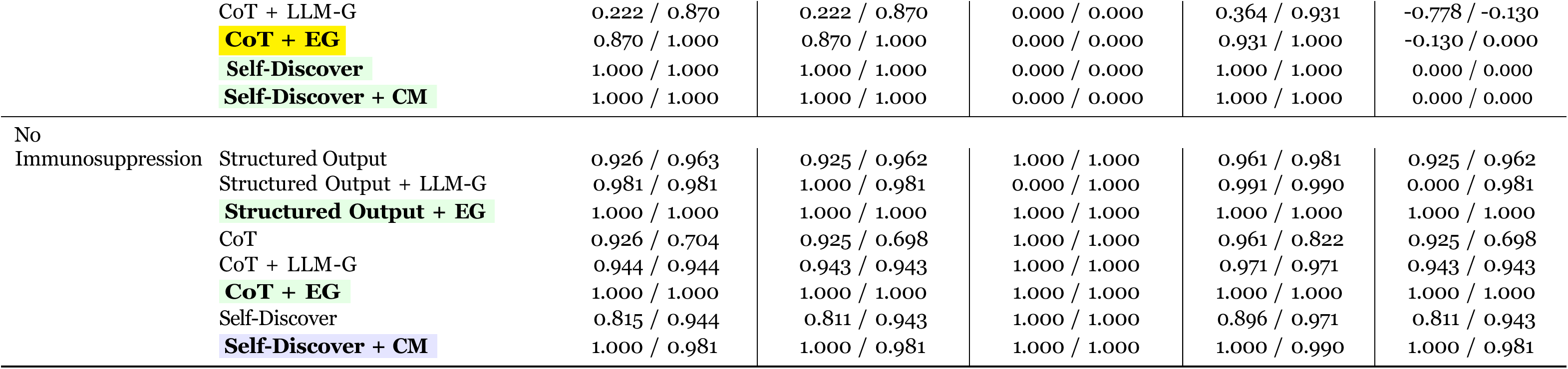
Comparison of Criterion-Level Performance Metrics for GPT-3.5 and GPT-4 on All Criteria Using Different Prompting Approaches. Abbreviations: Chain of Thought, CoT; Expert Guidance, EG; Custom Modules, CM; Accuracy, Acc; Sensitivity, Sens; Specificity, Spec; Micro F1, F1; Youden Index, YI; Micro F1, F1. The best approach by LLM measured using the YI: GPT-3.5, GPT-4, Both.

**Table C3.** Patient Level Performance Metrics Requiring all Criteria to be Met for Recommendation for Enrollment. Abbreviations: Acc, Accuracy; Sens, Sensitivity; Spec, Specificity; YI, Youden Index; LLM, Large Language Model; CoT, Chain of Thought; CM, Custom Modules. The best approach by LLM measured using the YI: GPT-3.5, GPT-4, Both.

| Criterion | Approach | Acc (GPT-3.5/4) | Sens (GPT-3.5/4) | Spec (GPT-3.5/4) | YI (GPT-3.5/4) |
| --- | --- | --- | --- | --- | --- |
| Overall | Structured | 0.537 / 0.630 | 0.000 / 0.200 | 1.000 / 1.000 | 0.000 / 0.200 |
|  | Structured + LLM-G | 0.500 / 0.537 | 0.000 / 0.000 | 0.931 / 1.000 | -0.069 / 0.000 |
|  | **Structured + EG** | 0.611 / 0.630 | 0.440 / 0.240 | 0.759 / 0.966 | 0.199 / 0.206 |
|  | CoT | 0.537 / 0.537 | 0.000 / 0.000 | 1.000 / 1.000 | 0.000 / 0.000 |
|  | CoT + LLM-G | 0.537 / 0.611 | 0.000 / 0.160 | 1.000 / 1.000 | 0.000 / 0.160 |
|  | **CoT + EG** | 0.593 / 0.648 | 0.120 / 0.240 | 1.000 / 1.000 | 0.120 / 0.240 |
|  | Self Discover | 0.537 / 0.574 | 0.000 / 0.080 | 1.000 / 1.000 | 0.000 / 0.080 |
|  | Self Discover + CM | 0.537 / 0.630 | 0.000 / 0.200 | 1.000 / 1.000 | 0.000 / 0.200 |
| Not Enrolled | **Structured** | 1.000 / 1.000 | 0.000 / 0.000 | 1.000 / 1.000 | 0.000 / 0.000 |
|  | **Structured + LLM-G** | 0.931 / 1.000 | 0.000 / 0.000 | 0.931 / 1.000 | -0.069 / 0.000 |
|  | Structured + EG | 0.759 / 0.966 | 0.000 / 0.000 | 0.759 / 0.966 | -0.241 / -0.034 |
|  | **CoT** | 1.000 / 1.000 | 0.000 / 0.000 | 1.000 / 1.000 | 0.000 / 0.000 |
|  | **CoT + LLM-G** | 1.000 / 1.000 | 0.000 / 0.000 | 1.000 / 1.000 | 0.000 / 0.000 |
|  | **CoT + EG** | 1.000 / 1.000 | 0.000 / 0.000 | 1.000 / 1.000 | 0.000 / 0.000 |
|  | **Self Discover** | 1.000 / 1.000 | 0.000 / 0.000 | 1.000 / 1.000 | 0.000 / 0.000 |
|  | **Self Discover + CM** | 1.000 / 1.000 | 0.000 / 0.000 | 1.000 / 1.000 | 0.000 / 0.000 |
| Enrolled | Structured | 0.000 / 0.200 | 0.000 / 0.200 | 0.000 / 0.000 | 0.000 / 0.200 |
|  | Structured + LLM-G | 0.000 / 0.000 | 0.000 / 0.000 | 0.000 / 0.000 | 0.000 / 0.000 |
|  | **Structured + EG** | 0.440 / 0.240 | 0.440 / 0.240 | 0.000 / 0.000 | 0.440 / 0.240 |
|  | **CoT** | 0.000 / 0.240 | 0.000 / 0.240 | 0.000 / 0.000 | 0.000 / 0.240 |
|  | CoT + LLM-G | 0.000 / 0.160 | 0.000 / 0.160 | 0.000 / 0.000 | 0.000 / 0.160 |
|  | **CoT + EG** | 0.120 / 0.240 | 0.120 / 0.240 | 0.000 / 0.000 | 0.120 / 0.240 |
|  | Self Discover | 0.000 / 0.080 | 0.000 / 0.080 | 0.000 / 0.000 | 0.000 / 0.080 |
|  | Self Discover + CM | 0.000 / 0.200 | 0.000 / 0.200 | 0.000 / 0.000 | 0.000 / 0.200 |

## Notes

### Competing Interest Statement

The authors have declared no competing interest.

### Funding Statement

This study did not receive any funding

### Author Declarations

Ethics committee/IRB of University of Texas Southwestern Medical Center gave ethical approval for this work

